# A randomized, placebo-controlled trial of a nasal spray solution containing broadly potent neutralizing antibodies against SARS-CoV-2 variants in healthy volunteers

**DOI:** 10.1101/2022.10.04.22280574

**Authors:** Thanarath Imsuwansri, Thitinan Jongthitinon, Niramon Pojdoung, Nuntana Meesiripan, Siriwan Sakarin, Chatikorn Boonkrai, Tossapon Wongtangprasert, Tanapati Phakham, Thittaya Audomsun, Chadaporn Attakitbancha, Pijitra Saelao, Phijitra Muanwien, Maoxin Tim Tian, Songsak Tongchusak, Bhrus Sangruji, Dhammika Leshan Wannigama, Chenphop Sawangmake, Watchareewan Rodprasert, Quynh Dang Le, Steven Dwi Purbantoro, Kananuch Vasuntrarak, Sirirat Nantavisai, Supakit Sirilak, Ballang Uppapong, Sompong Sapsutthipas, Sakalin Trisiriwanich, Thitiporn Somporn, Asmah Usoo, Natthakarn Mingngamsup, Supaporn Phumiamorn, Porawan Aumklad, Kwanputtha Arunprasert, Prasopchai Patrojanasophon, Praneet Opanasopit, Norapath Pesirikan, Ladda Nitisaporn, Jesada Pitchayakorn, Thana Narkthong, Bancha Mahong, Kumchol Chaiyo, Kanjana Srisuthisamphan, Ratchanont Viriyakitkosol, Songklot Aeumjaturapat, Anan Jongkaewwattana, Sakarn Bunnag, Trairak Pisitkun

**Affiliations:** National Cancer Institute, Department of Medical Services, Ministry of Public Health, Bangkok, Thailand; Center of Excellence in Systems Biology, Faculty of Medicine, Chulalongkorn University, Bangkok, Thailand; The Excellence Chulalongkorn Comprehensive Cancer Center, King Chulalongkorn Memorial Hospital, Bangkok, Thailand; School of Arts and Sciences, Tufts University, Medford, United States of America; Department of Microbiology, Faculty of Medicine, Chulalongkorn University, Bangkok, Thailand; School of Medicine, Faculty of Health and Medical Sciences, The University of Western Australia, Western Australia, Australia; Department of Pharmacology, Faculty of Veterinary Science, Chulalongkorn University, Bangkok, Thailand; Veterinary Stem Cell and Bioengineering Innovation Center (VSCBIC), Faculty of Veterinary Science, Chulalongkorn University, Bangkok, Thailand; Veterinary Stem Cell and Bioengineering Research Unit, Faculty of Veterinary Science, Chulalongkorn University, Bangkok, Thailand; Academic Affairs, Faculty of Veterinary Science, Chulalongkorn University, Bangkok, Thailand; Department of Medical Sciences, Ministry of Public Health, Nonthaburi, Thailand; Institute of Biological Products, Department of Medical Sciences, Ministry of Public Health, Nonthaburi, Thailand; Faculty of Pharmacy, Silpakorn University, Nakhon Pathom, Thailand; The Government Pharmaceutical Organization, Bangkok, Thailand; Virology and Cell Technology Research Team, National Center for Genetic Engineering and Biotechnology (BIOTEC), National Science and Technology Development Agency (NSTDA), Pathumthani, Thailand; Otolaryngology Department, King Chulalongkorn Memorial Hospital, Bangkok, Thailand

**Keywords:** ACE2, mucosal immunity, hypromellose, pseudovirus neutralization, medical device, vaccine

## Abstract

Successful COVID-19 prevention requires additional measures beyond vaccination, social distancing, and masking. A nasal spray solution containing human IgG1 antibodies against SARS-CoV-2 (COVITRAP™) was developed to strengthen other COVID-19 preventive arsenals. Here, we evaluated its pseudovirus neutralization potencies, preclinical and clinical safety profiles, and intranasal SARS-CoV-2 inhibitory effects in healthy volunteers (NCT05358873). COVITRAP™ exhibited broadly potent neutralizing activities against SARS-CoV-2 with PVNT_50_ values ranging from 0.0035 to 3.1997 μg/ml for the following variants of concern (ranked from lowest to highest): Alpha, Beta, Gamma, Ancestral, Delta, Omicron BA.1, Omicron BA.2, Omicron BA.4/5, and Omicron BA.2.75. It demonstrated satisfactory preclinical safety profiles based on evaluations of *in vitro* cytotoxicity, skin sensitization, intracutaneous reactivity, and systemic toxicity. Its intranasal administration in rats did not yield any detected circulatory levels of the human IgG1 anti-SARS-CoV-2 antibodies at any time point during the 120 hours of follow-up. A double-blind, randomized, placebo-controlled trial (RCT) was conducted on 36 healthy volunteers who received either COVITRAP™ or a normal saline nasal spray at a 3:1 ratio. Safety of the thrice-daily intranasal administration for 7 days was assessed using nasal sinuscopy, adverse event recording, and self-reporting questionnaires. COVITRAP™ was well tolerated, with no significant adverse effects in healthy volunteers for the entire 14 days of the study. The intranasal SARS-CoV-2 inhibitory effects of COVITRAP™ were evaluated in nasal fluids taken from volunteers pre- and post-administration using a SARS-CoV-2 surrogate virus neutralization test. SARS-CoV-2 inhibitory effects in nasal fluids collected immediately or six hours after COVITRAP™ application were significantly increased from baseline for all three variants tested, including Ancestral, Delta, and Omicron BA.2. In conclusion, COVITRAP™ was safe for intranasal use in humans to provide SARS-CoV-2 inhibitory effects in nasal fluids that lasted at least six hours. Therefore, COVITRAP™ can be considered an integral instrument for COVID-19 prevention.

## INTRODUCTION

The enduring waves of SARS-CoV-2 breakthrough infections create a global impediment that requires additional measures beyond vaccination to mitigate this perpetual situation. SARS-CoV-2 is an RNA virus with the characteristic of multiple spike glycoproteins on its envelope^1^. Through airborne transmission, the nasopharyngeal epithelium is SARS-CoV-2’s primary portal, which incubates the virus to a high viral load for shedding and dissemination^2,3^. The receptor-binding domain (RBD) on SARS-CoV-2 spike glycoproteins specifically binds angiotensin-converting enzyme 2 (ACE2) expressed on the plasma membrane of target cells, setting off a cell entry cascade of the virus^4^. The local defense system at the nasopharyngeal mucosa, especially via antibody-mediated immunity that rapidly interferes with RBD-ACE2 engagement, is thus regarded as the genuine instrument for COVID-19 prevention^5-7^. However, after systemic vaccination, the neutralizing antibody levels in the nasopharyngeal mucosa naturally decline and are typically insufficient to prevent SARS-CoV-2 breakthrough infections in the long term. However, it is impractical to repeatedly boost systemic vaccines to maintain the protective level of mucosal immunity at all times. Therefore, an innovative approach is needed for this unprecedented situation. Recently, strategies to bolster mucosal immunity using an active or passive route via intranasal administration of vaccines or antibodies, respectively, have gained critical momentum^5,8-12^.

COVITRAP™ is a nasal spray solution containing human IgG1 antibodies. It has been approved by the Thai FDA as an innovative medical device platform (Class 4) to support mucosal immunity against SARS-CoV-2 infections via a dual mechanism of action through antibody-mediated specific inhibition coupled with a steric barrier (Figure 1). Human IgG1 antibodies included in the COVITRAP™ platform are monoclonal antibodies with broadly potent neutralizing activities against SARS-CoV-2 spike glycoproteins screened from elite responders who have fully recovered from COVID-19. COVITRAP™ is strategically formulated to allow a timely modification of the antibody component to react to the potential immune escape of future SARS-CoV-2 variants. In addition to human IgG1 antibodies, a mucoadhesive cellulose derivative, hypromellose (HPMC), is another key ingredient of COVITRAP™ that forms a steric barrier on nasopharyngeal mucosa to prevent SARS-CoV-2 from entering target cells. This study aims to evaluate COVITRAP™’s pseudovirus neutralization potencies, preclinical and clinical safety profiles, and intranasal SARS-CoV-2 inhibitory effects in healthy volunteers in accordance with the ICH-GCP guidelines.

**Figure 1.**
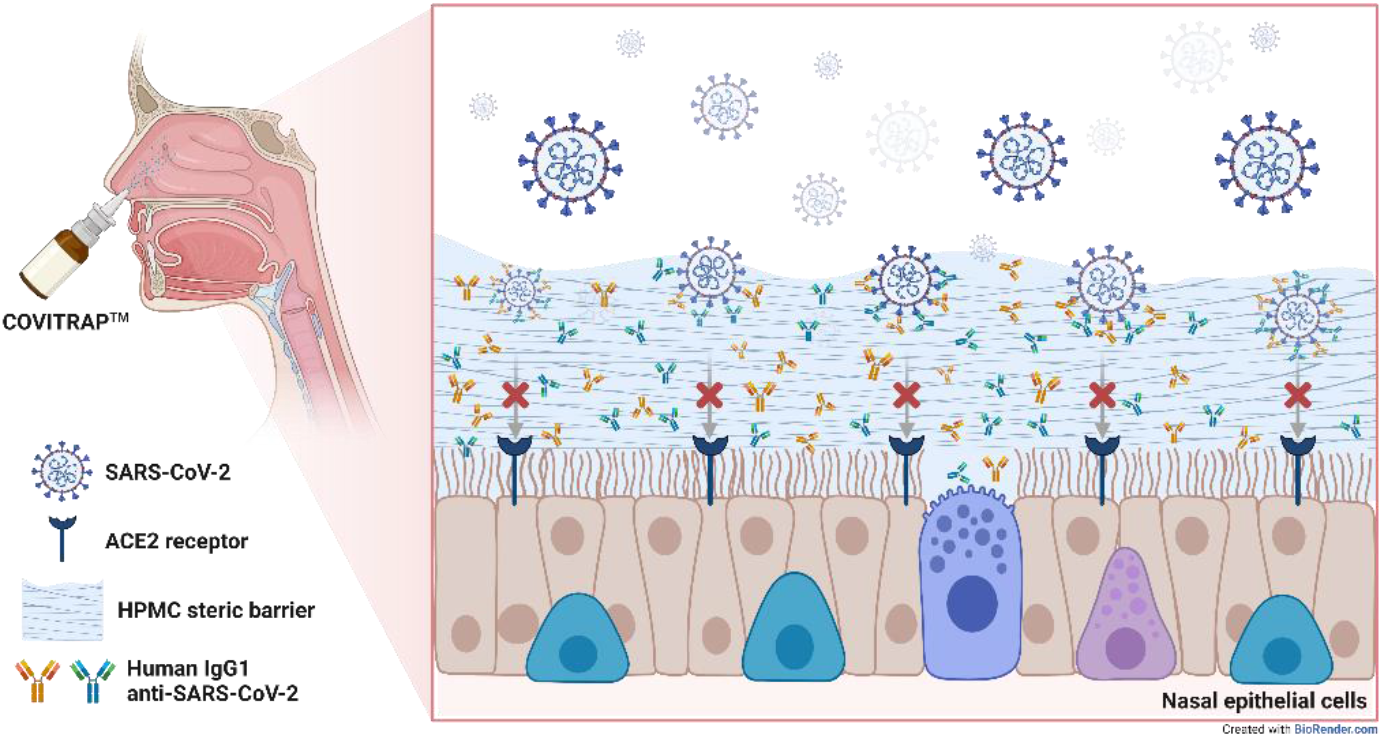
Mechanism of action of COVITRAP™ nasal spray

## RESULTS

### Pseudovirus neutralization potencies

Pseudovirus microneutralization assays were performed as previously described^13,14^ to assess the neutralization potency of COVITRAP™ against various SARS-CoV-2 pseudoviruses, i.e., Ancestral, Alpha, Delta, Omicron BA.1, Omicron BA.2, Omicron BA.4/5, and Omicron BA.2.75. The results revealed that COVITRAP™ potently neutralized all mentioned pseudoviruses with 50% pseudovirus neutralization titers (PVNT_50_) ranging from 0.0035 to 3.1997 μg/ml (Table 1). It should be noted that the magnitude of neutralization against Omicron BA.2.75, which is among the most immune-evasive SARS-CoV-2 variants, appears to be less potent than that of other variants. However, the concentration of the antibody cocktail in COVITRAP™ is still 78.13-fold higher than Omicron BA.2.75’s PVNT_50_.

**Table 1.** Pseudovirus neutralization potencies of COVITRAP™

| SARS-CoV-2 variants | PVNT<sub>50</sub> (µg/ml) |
| --- | --- |
| Ancestral | 0.0092 |
| Alpha | 0.0035 |
| Beta | 0.0044 |
| Gamma | 0.0055 |
| Delta | 0.0117 |
| Omicron BA.1 | 0.0219 |
| Omicron BA.2 | 0.0135 |
| Omicron BA.4/5 | 0.1919 |
| Omicron BA.2.75 | 3.1997 |

### Biocompatibility

The results of all biocompatibility studies are summarized in Table 2 and Supplemental Data S1. An *in vitro* cytotoxicity study by the direct contact method demonstrated that COVITRAP™ was noncytotoxic to the Balb/c 3T3 cell line. A skin sensitization study in guinea pigs revealed that COVITRAP™ was considered a nonsensitizer. An irritation study in New Zealand white rabbits by an intracutaneous reactivity test demonstrated that COVITRAP™ was deemed nonreactive to rabbits. Acute systemic toxicity results indicated that COVITRAP™ did not induce any systemic toxicity in Swiss albino mice. Last, 28-day subacute systemic toxicity study data showed no mortality/morbidity or any other clinical signs of toxicity during the study period.

**Table 2.**
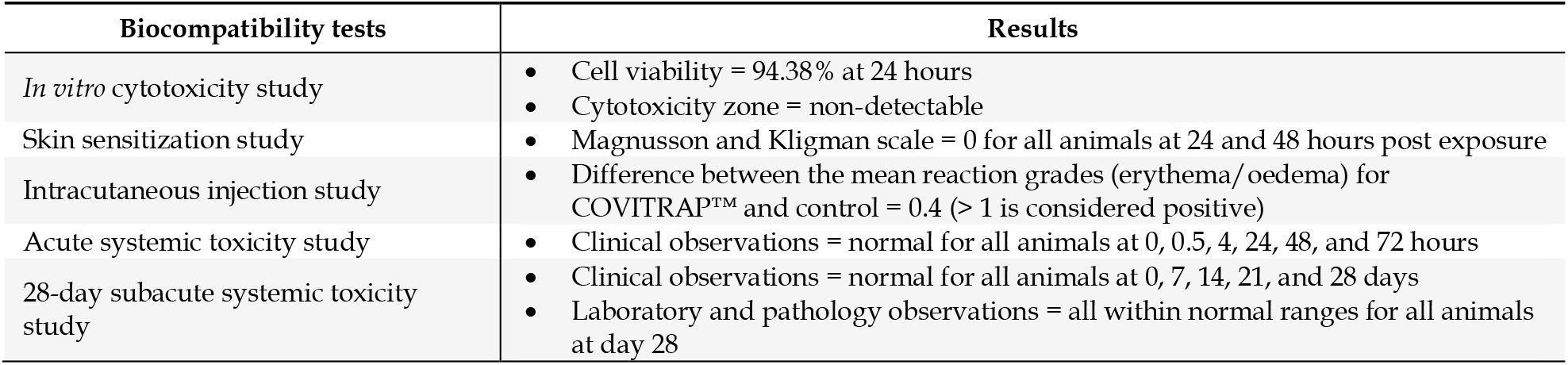
Biocompatibility assessment of COVITRAP™

### Circulatory levels of human IgG1 anti-SARS-CoV-2 antibodies after intranasal application of COVITRAP™

Quantitative measurement of human IgG1 anti-SARS-CoV-2 antibodies after intranasal application of COVITRAP™ was determined in rats. A single dose of COVITRAP™ (20 μg/kg) was intranasally applied to 10-week-old female Sprague-Dawley rats (n=13). This dose was calculated based on the intended single-use amount in humans (2 μg/kg) multiplied by a human-to-rat conversion factor of 10^15^. Three rats without any intervention were used as controls. Rat serum was collected at 1, 4, 8, 24, 48, 72, 96, and 120 hours post-administration. Human IgG1 anti-SARS-CoV-2 antibodies were not detected by ELISA in the bloodstream of rats at any time point during the 120 hours post intranasal administration (limit of quantitation = 0.165 ng/ml).

### Clinical study of COVITRAP™

Figure 2 shows a flow chart of this study. Thirty-eight healthy volunteers were enrolled. However, two of them were excluded due to nasal polyps. Thirty-six participants were randomly assigned in a 1:3 ratio to receive a placebo or COVITRAP™. The characteristics of all participants enrolled in the study are summarized in Table 3.

**Table 3.** Baseline characteristics of participants in the study

| Characteristics | Placebo<br>(n=9) | COVITRAP™<br>(n=27) | Total<br>(n=36) |
| --- | --- | --- | --- |
| Age (median, range) | 30 (24-45) | 32 (24-45) | 32 (24-45) |
| Gender; n (%) |  |  |  |
| Male | 6 (66.66) | 12 (44.44) | 18 (50.00) |
| Female | 3 (33.33) | 15 (55.57) | 18 (50.00) |
| Average height, cm | 166.56 | 165.41 | 165.69 |
| Average weight, kg | 67.29 | 63.36 | 64.34 |
| Average BMI, kg/m<sup>2</sup> | 24.05 | 23.06 | 23.31 |
| Average blood pressure (systolic/ diastolic), mmHg | 127.33/73.22 | 121.65/72.12 | 123.11/72.40 |
| Average heart rate, beats per minute | 79.56 | 81.69 | 81.14 |
| Average respiration rate, breath per minute | 19.22 | 18.80 | 18.91 |
| Average spO<sub>2</sub>, % | 99 | 98.5 | 98.75 |
| Fully vaccinated against covid-19, % | 100 | 100 | 100 |

**Figure 2.**
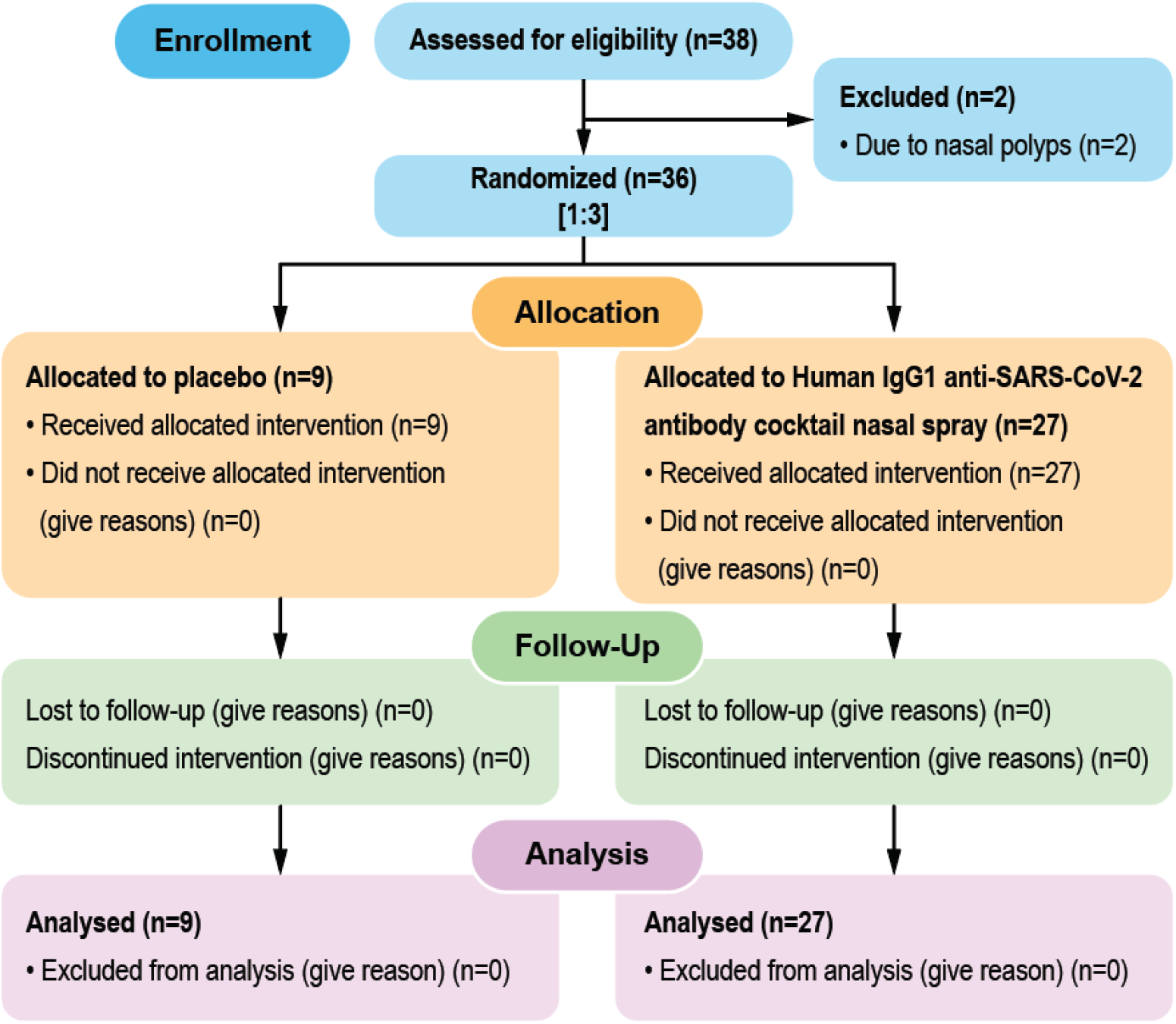
Study flow chart.

### Safety and tolerability of COVITRAP™

To evaluate the safety and tolerability of COVITRAP™, we used objective (nasal sinuscopy) and subjective (SNOT-22 and TNSS questionnaires) assessments. Participants sprayed 2 puffs of the study products into each nostril thrice daily at 8 am, 2 pm, and 8 pm for 7 days and were then followed up for another 7 days. Nasal sinuscopy was performed on all participants on days 0, 7, and 14. Nasal sinuscopy findings were evaluated using the modified Lund-Kennedy endoscopic scoring system. Following this scoring system, we did not find any changes in nasal mucosa appearance or any signs of inflammation in either the COVITRAP™ or placebo group at any given time point (Figure 3 and Table 4). Supplemental Data S2 shows nasal sinuscopy images displayed in random order of participants.

**Table 4.** Safety assessment via nasal sinuscopy

| Physical examination via nasal sinuscopy | Placebo<br>(n=9) |  |  | COVITRAP™<br>(n=27) |  |  |
| --- | --- | --- | --- | --- | --- | --- |
| *Day* | 0 | 7 | 14 | 0 | 7 | 14 |
| *Total modified Lund-Kennedy endoscopic score (mean ± SE)* | 0 ± 0 | 0 ± 0 | 0 ± 0 | 0 ± 0 | 0 ± 0 | 0 ± 0 |
| *Two-tailed t-test* | Not statistically significant |  |  |  |  |  |

**Figure 3.**
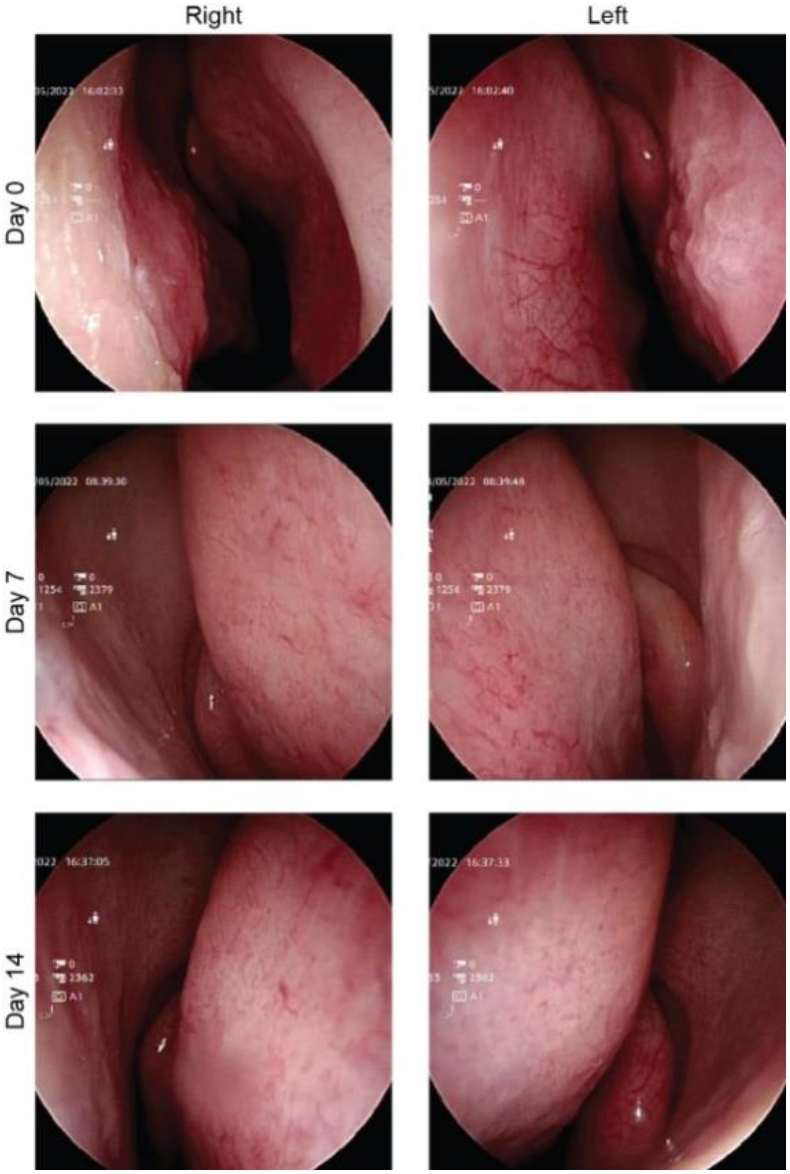
Representative nasal sinuscopy images of participants who received COVITRAP™ on days 0, 7, and 14.

SNOT-22 and TNSS questionnaires were used to evaluate nasal symptoms throughout the two weeks of the study period. These questionnaires have been validated for patient-reported outcomes of chronic rhinosinusitis, allergic rhinitis, and other sinonasal outcomes^16,17^. For both questionnaires, each participant scored the severity of each symptom daily. Fourteen-day accumulative severity scores for each symptom were compared between the COVITRAP™ and placebo groups. The results showed that for both questionnaires, the highest reported score was 0 (no problem or none) for every symptom in both groups. For some symptoms, especially runny nose/rhinorrhea, a score of 1 (SNOT-22: a very mild problem and TNSS: mild) was reported at a low percentage in the COVITRAP™ group; however, these mild nasal symptoms were recovered without any medical treatments in all cases. The runny nose symptom is likely a result of the slightly viscous HPMC-based solution of COVITRAP™. HPMC helps extend the retention time of the antibodies in the nasal cavity by reducing mucociliary clearance and might explain this nasal symptom. Overall, both groups had no substantial difference in sinonasal symptoms (Tables 5 and 6).

**Table 5.**
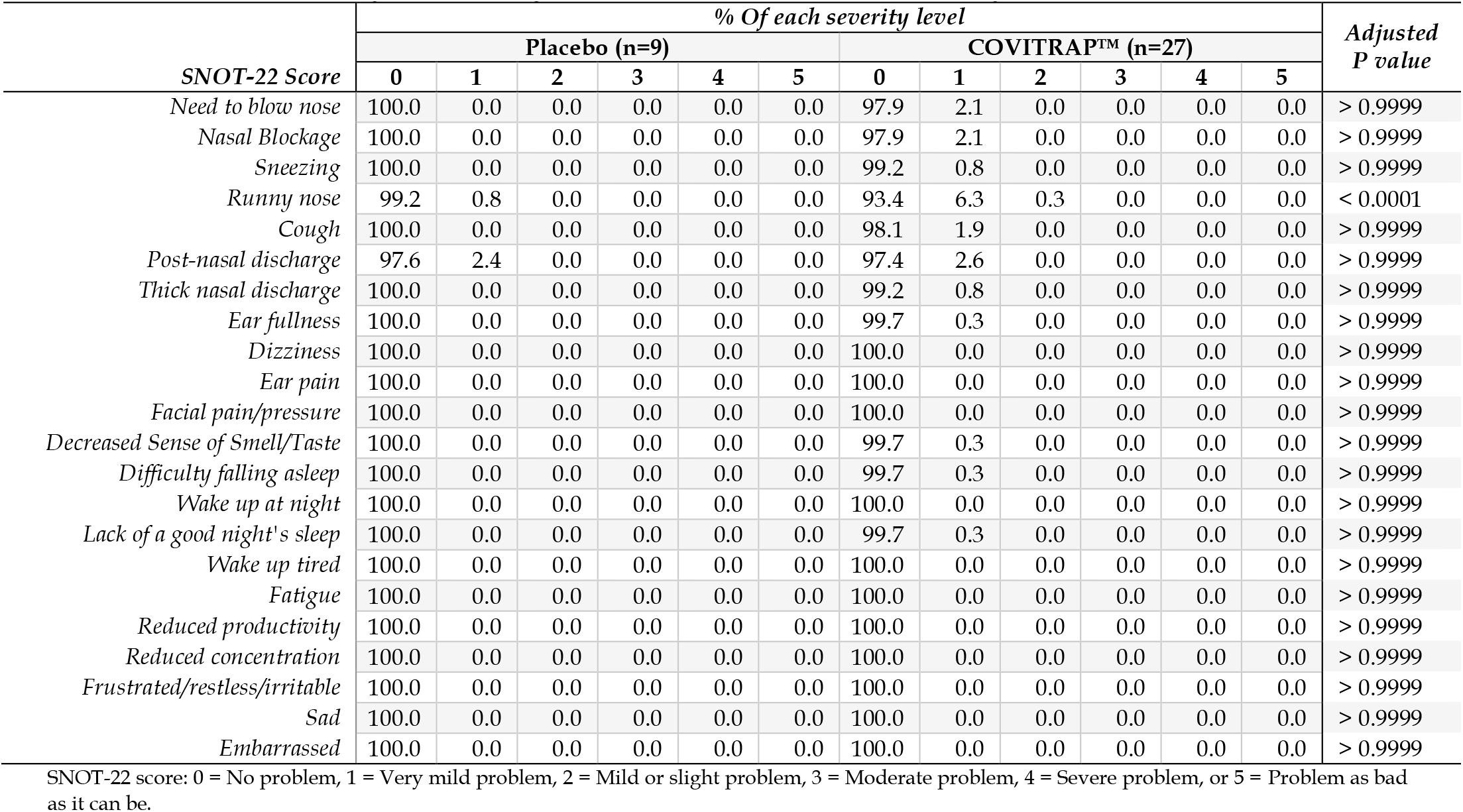
Self-reported symptoms by SNOT-22 questionnaire (14-day accumulative events)

**Table 6.** Self-reported symptoms by TNSS questionnaire (14-day accumulative events)

| TNSS score | % Of each severity level |  |  |  |  |  |  |  | Adjusted<br>P value |
| --- | --- | --- | --- | --- | --- | --- | --- | --- | --- |
|  | Placebo (n=9) |  |  |  | COVITRAP™ (n=27) |  |  |  |  |
|  | 0 | 1 | 2 | 3 | 0 | 1 | 2 | 3 |  |
| Rhinorrhea | 100.0 | 0.0 | 0.0 | 0.0 | 94.2 | 5.6 | 0.3 | 0.0 | 0.0005 |
| Nasal congestion | 100.0 | 0.0 | 0.0 | 0.0 | 98.7 | 1.3 | 0.0 | 0.0 | > 0.9999 |
| Nasal itching | 100.0 | 0.0 | 0.0 | 0.0 | 98.9 | 1.1 | 0.0 | 0.0 | > 0.9999 |
| Sneezing | 100.0 | 0.0 | 0.0 | 0.0 | 98.7 | 1.3 | 0.0 | 0.0 | > 0.9999 |

TNSS score: 0 = None, 1 = Mild, 2 = Moderate, or 3 = Severe

Additionally, treatment-emergent adverse events (TEAEs) were evaluated over 14 days of the study. No adverse events were reported from participants in either group (Table 7). Collectively, all assessments indicated that COVITRAP™ was well tolerated, with no significant adverse effects in healthy volunteers.

**Table 7.** Self-reported treatment emergent adverse events (TEAEs)

| Adverse events | No. of TEAEs |  |
| --- | --- | --- |
|  | Placebo<br>(n=9) | COVITRAP™<br>(n=27) |
| Fatal (resulted in death) | 0 | 0 |
| A life-threatening occurrence | 0 | 0 |
| Requires inpatient hospitalization or prolongation of existing hospitalization | 0 | 0 |
| Results in persistent or significant disability/incapacity | 0 | 0 |
| Results in congenital anomaly/birth defect | 0 | 0 |
| A significant medical incident that, based upon appropriate medical judgment, may jeopardize the subject, and require medical or surgical intervention to prevent one of the outcomes listed above. | 0 | 0 |

### Intranasal SARS-CoV-2 inhibitory effects of COVITRAP™

Nasal fluid was collected by swabbing from both nostrils before and immediately or 6 hours after the study product application in both the COVITRAP™ and placebo groups (see Figure 4A). A SARS-CoV-2 surrogate virus neutralization test was utilized to determine SARS-CoV-2 inhibitory effects in the collected nasal fluids.

**Figure 4.**
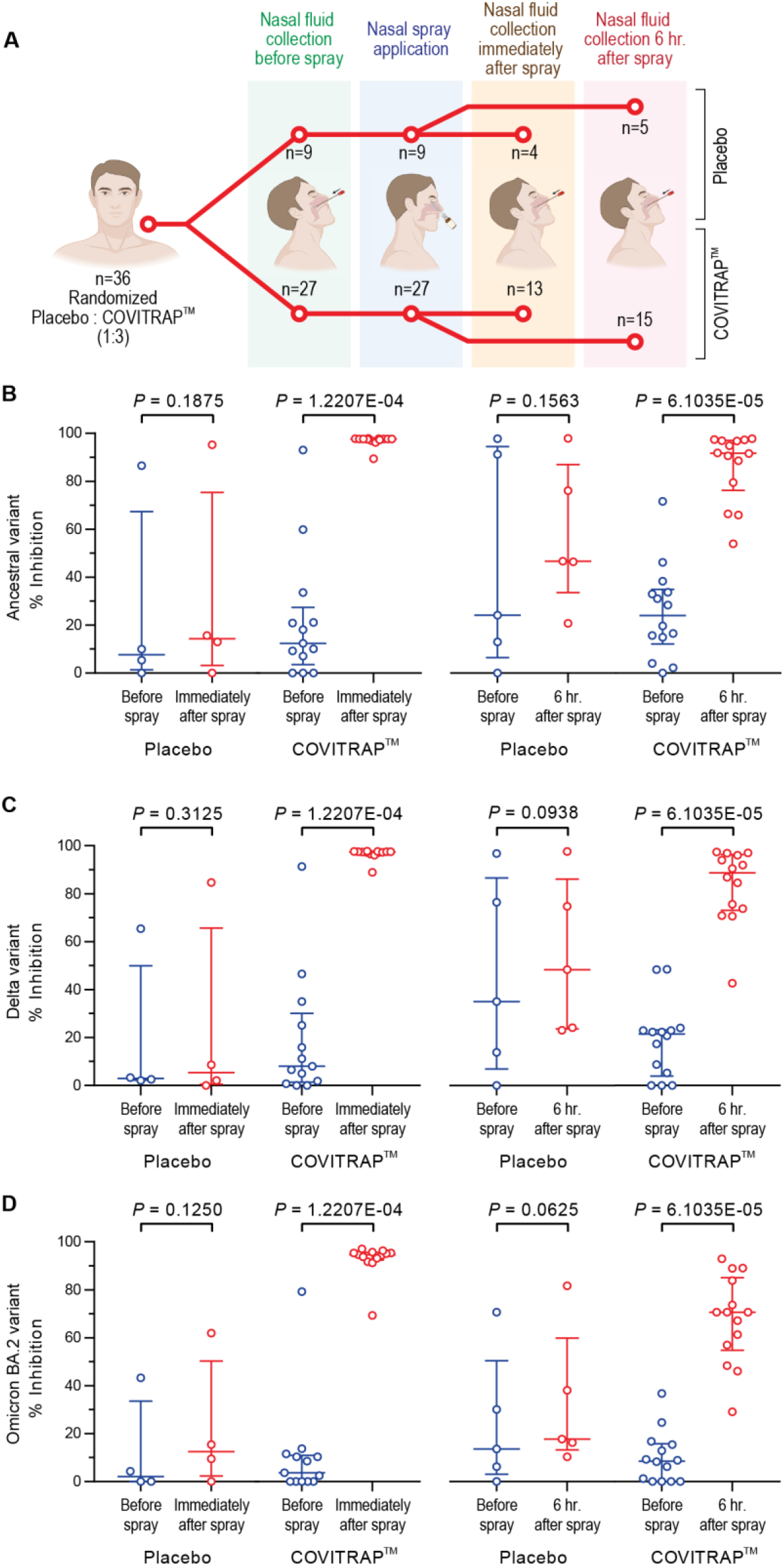
Intranasal SARS-CoV-2 inhibitory effects of COVITRAP™. (A) Study design. % Inhibition against Ancestral (B), Delta (C), and Omicron BA.2 (D) variants from nasal fluids swabbed before and after the study product application. Data are presented in IQR ± 25th-75th percentile.

The percent inhibition against the ancestral variant of the nasal fluid from the COVITRAP™ group was significantly increased from baseline at both time points (median percent inhibition of baseline vs. immediate time point: 12.41% vs. 97.58%; P-value = 1.2207E-4 and median percent inhibition of baseline vs. 6 hours time point: 24.03% vs. 91.72%; P-value = 6.1035E-05), see Figure 4B. Similarly, for the Delta and Omicron BA.2 variants (Figures 4C and 4D), the nasal fluid from the COVITRAP™ group showed a significant increase in the percent inhibition from the baseline at the immediate time point for both variants (median percent inhibition of baseline vs. immediate time point for Delta and Omicron BA.2 variants: 8.02% vs. 97.44%; P-value = 1.2207E-4 and 3.64% vs. 94.65%; P-value = 1.2207E-4). Likewise, the results at the 6-hour time point still demonstrated a significant increase in the percent inhibition from baseline for both variants (median percent inhibition of baseline vs. 6-hour time point for Delta and Omicron BA.2 variants: 21.53% vs. 88.67%; P-value = 6.1035E-05 and 8.59% vs. 70.60%; P-value = 6.1035E-05). In contrast, the nasal fluid at both time points from the placebo group did not show a significant difference in the percent inhibition from the baseline for all three variants tested. Supplemental Data S3 contains a detailed statistical report of the intranasal SARS-CoV-2 inhibitory effects of COVITRAP™.

## DISCUSSION

COVITRAP™ is a medical device innovated to support mucosal immunity via a dual mechanism of action in which a broadly potent neutralizing human IgG1 anti-SARS-CoV-2 monoclonal antibody cocktail produces inhibitory effects against multiple variants of concern (VOCs) in nasal fluid, and a steric barrier-forming agent, HPMC, fortifies the mucus layer. COVITRAP™ exhibited broadly neutralizing activities against SARS-CoV-2 pseudoviruses of Ancestral, Alpha, Delta, Omicron BA.1, Omicron BA.2, Omicron BA.4/5, and Omicron BA.2.75 variants. The preclinical studies following the ISO 10993 standards of medical devices showed good biocompatibility based on cytotoxicity, skin sensitization, and intracutaneous reactivity evaluations as well as satisfactory safety profiles by both acute and subacute systemic toxicity investigations. Intranasal administration of COVITRAP™ did not result in any detection of human IgG1 anti-SARS-CoV-2 antibodies in the bloodstream of rats at any time point during the 120 hours of follow-up. This finding agrees with the knowledge that the nasal epithelial barrier only allows the passage of molecules smaller than 1,000 Da^18^; thus, human IgG1’s molecular mass of 146,000 Da^19^ clearly prohibits the systemic distribution of the human IgG1 antibodies in COVITRAP™. The randomized, placebo-controlled trial of intranasal administration of COVITRAP™ in 36 healthy participants revealed that COVITRAP™ was well tolerated, without any changes in nasal mucosa appearance, any signs of inflammation, or any treatment-emergent adverse events for the entire 14 days of the study.

Recently, the antibody neutralization level was shown to be highly predictive of immune protection and correlate with the risk for COVID-19^20,21^. Therefore, we exploited the neutralizing antibody level in nasal fluids as a proxy for the COVID-19 protection ability of COVITRAP™ in this study. The neutralizing antibody level in nasal fluids producing intranasal SARS-CoV-2 inhibitory effects correlates with the potential protective effects against omicron infection^7^. We found a significant increase in intranasal SARS-CoV-2 inhibitory effects after COVITRAP™ application compared with baseline for all three variants tested, including Ancestral, Delta, and Omicron BA.2. Immediately after COVITRAP™ application, SARS-CoV-2 inhibitory effects in nasal fluids were increased to ≥ 91.69%. These intranasal effects remained significantly increased at 6 hours after the application (≥ 70.60%). HPMC in COVITRAP™ acts as a viscosity-inducing agent that helps extend the retention time of the antibodies in the nasal cavity by reducing mucociliary clearance^22^. Further study is needed to define the precise duration of the intranasal SARS-CoV-2 inhibitory effects of COVITRAP™.

Collectively, COVITRAP™ can safely and effectively support mucosal immunity at the point of entry of the virus, making it an essential and complementary tool in our preexisting COVID-19 prevention arsenals. Nevertheless, a large-scale efficacy trial measuring COVID-19 incidence will be required to demonstrate the efficacy of COVID-19 prevention by COVITRAP™.

## MATERIALS AND METHODS

### Study product

COVITRAP™ is an HPMC-based nasal spray solution containing human IgG1 anti-SARS-CoV-2 monoclonal antibodies, clones 1D1 and 3D2 [US application No. US 63/248,115 and PCT/TH2022/000037] at 0.25 mg/ml. The concentration of the antibody cocktail in COVITRAP™ was established at 833.33 times the 99% plaque reduction neutralization test (PRNT_99_) value against the SARS-CoV-2 delta variant [data from the patent application].

### Pseudovirus-based neutralization assay

Lentiviral pseudoviruses containing SARS-CoV-2 spike were produced with slight modifications, as previously described by Di Genova et al.^23^. To generate pseudoviruses, a lentivirus backbone expressing a firefly luciferase reporter gene (pCSFLW), an expression plasmid expressing HIV-1 structural/regulatory proteins (pCMVR8.91), and pCAGGS expressing spike constructs were used. Unless otherwise noted, HEK293T/17 producing cells were seeded in 6-well plates 24 hours before transfection with 600 ng pCMVR8.91, 600 ng pCSFLW, and 500 ng pCAGGS spike in OptiMEM containing 10 μl polyethylenimine (PEI). Transfected cells were incubated at 37°C with 5% CO_2_. Twelve hours after transfection, the cells were washed and grown in DMEM containing 10% FBS. Pooled supernatants containing pseudoviruses were collected 72 hours after transfection, centrifuged at 1,500×*g* for 10 minutes at 4°C to remove cell debris, aliquoted, and kept at 80°C.

To assess the neutralizing activities of human IgG1 antibodies in COVITRAP™, a two-fold serial dilution of the nasal spray solution was conducted in a culture medium starting at a ratio of 1:40 (high-glucose DMEM without FBS). In a 96-well culture plate, the diluted samples were mixed with pseudoviruses bearing the SARS-CoV-2 spike of interest at a 1:1 v/v ratio. The input pseudovirus was adjusted to 1×10^5^ RLU per well. The antibody-pseudovirus mixture was then incubated at 37°C for 1 hour. Cell suspensions of HEK293T-ACE-2 cells pretransfected with pCAGGS expressing human TMPRSS2 (2×10^4^ cells/ml) were then seeded into each well of CulturPlate™ microplates (PerkinElmer). Finally, plates were incubated at 37°C for 48 hours, and neutralizing activities were detected by measuring luciferase activity, as previously described^24^.

### Biocompatibility testing

The biocompatibility of COVITRAP™ was evaluated by the following 5 assessments conforming to the standards of medical devices (ISO 10993-5:2009, ISO 10993-10:2021, ISO 10993 Part 23: 2021, and ISO 10993-11:2017): 1) *in vitro* cytotoxicity using the direct contact method, 2) skin sensitization using the guinea pig maximization test, 3) intracutaneous reactivity potentials in New Zealand white rabbits, 4) acute systemic toxicity study via intranasal administration in mice, and 5) 28-day subacute systemic toxicity study via oral administration in rats (see details in Supplemental Methods).

### Enzyme-linked immunosorbent assay (ELISA)

ELISA was employed to quantify human IgG1 anti-SARS-CoV-2 antibody levels in the circulation after intranasal application of COVITRAP™ in rats. In brief, rat serum samples diluted in 3% BSA in PBS buffer at a 1:10 dilution were added (100 μl/well) to an ELISA plate coated with 100 ng/well of Delta-variant RBD proteins (Sino Biological, 40592-V08H90). Human IgG1 anti-SARS-CoV-2 antibodies were detected with goat anti-human IgG Fcγ-HRP antibody (Jackson Immuno Research, 109-005-098) diluted 1:2,000 in 3% BSA in PBS buffer. The SIGMA*FAST*™ OPD (Sigma-Aldrich, P9187) substrate solution was used, and the reaction was stopped by adding 1 M H_2_SO_4_. The absorbance was measured at 492 nm by a Cytation™ 5 cell imaging multi-mode reader (BioTek).

### Clinical study design

This study was designed as a single-center, double-blind, randomized, placebo-controlled trial to evaluate the safety, tolerability, and clinical performance of COVITRAP™ in healthy volunteers. The protocol was registered with ClinicalTrials.gov (NCT05358873).

### Ethical considerations

This study was approved by the Ethics Committee of the Department of Medical Services, Ministry of Public Health, Thailand (Approval No. 0001/2565). All procedures were performed following the principles of the Declaration of Helsinki and the ICH-GCP guidelines. All participants provided written informed consent before the commencement of the study and voluntarily participated in this clinical trial.

### Participants

We calculated the sample size based on previous recommendations^25,26^ using Z statistics to assess the product’s safety, tolerability, and performance. We set a power of 69.0% to detect an effect size (E) of 0.5 with a threshold probability of rejecting the null hypothesis (α) of 5% (two-tailed). Furthermore, it was assumed that the data had a 20% probability of not rejecting the null hypothesis under the alternative hypothesis (β) and a standard deviation of change (SΔ) of the outcome of 1. The sample size (n) was calculated per the following formula.

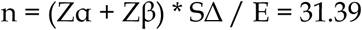

To ensure an adequate sample size in case participants dropped out, we added 4 more participants to attain the total number of 36 participants. Healthy volunteers interested in participating in the study were screened according to the inclusion and exclusion criteria described on ClinicalTrials.gov (NCT05358873).

All volunteers were randomly assigned in a 1:3 ratio into 2 groups: placebo (n=9) and COVITRAP™ (n=27), as shown in Figure 1.

### Product application

COVITRAP™ and normal saline solution (placebo) were produced, packaged, and labeled with a double-blind, randomized code by the Government Pharmaceutical Organization (GPO), Ministry of Health, Thailand. Each participant was randomly assigned to receive either COVITRAP™ or a placebo. Site staff gave participants instructions on the study product application, storage, and return. Participants sprayed 2 puffs (100 μl per puff) of the study products into each nostril thrice daily at 8 am, 2 pm, and 8 pm for 7 days. The volume and frequency of the study product application were based on reports of other nasal spray products containing anti-SARS-CoV-2 antibodies^27,28^. Any nasal products other than the study products were prohibited during the study period.

### Safety evaluation

Safety was assessed based on nasal sinuscopy examination, treatment-emergent adverse events (TEAEs), and Sino-Nasal Outcome Test-22 (SNOT-22) and Total Nasal Symptom Score (TNSS) questionnaires^16,29^. Nasal sinuscopy was performed on all participants using the Olympus ENF-V4 video rhinolaryngoscope on days 0, 7, and 14. Nasal sinuscopy findings were evaluated by an otorhinolaryngologist using the modified Lund-Kennedy endoscopic scoring system. Participants were asked to complete the SNOT-22 and TNSS questionnaires daily until the end of the study (Day 14).

### SARS-CoV-2 surrogate virus neutralization test

SARS-CoV-2 inhibitory effects in nasal fluid specimens against the ancestral, Delta, and Omicron BA.2 HRP-conjugated RBD proteins were determined using a SARS-CoV-2 surrogate virus neutralization test (cPass™ SARS-CoV-2 Neutralization Antibody Detection Kit, GenScript; A02087, Z03614, and Z03741). Nasal fluid specimens were diluted ∼10-fold in sample dilution buffer, and then the SARS-CoV-2 inhibitory effects were measured according to the instruction manual and reported as % inhibition against SARS-CoV-2.

### Statistical analysis

For the safety assessments of COVITRAP™, an unpaired two-tailed Mann–Whitney U test was used, and Dunn’s test was applied for multiple testing corrections. For the assessments of intranasal SARS-CoV-2 inhibitory effects of COVITRAP™, the difference in % inhibition before and after the study product applications was compared using a one-tailed Wilcoxon signed-rank test.

## Data Availability

All data produced in the present work are contained in the manuscript.

## DATA AVAILABILITY

All data generated or analyzed during this study are included in this published article.

## ACKNOWLEDGMENTS

This study was sponsored by HIBIOCY Co., Ltd., and funded by the Program Management Unit Competitiveness (PMU-C) [Grant No. C10F650051], Ministry of Higher Education, Science, Research and Innovation, Thailand. Data collection and analysis were supported by Ever Medical Technology Co., Ltd., and illustrations were created with *BioRender*.*com*.

## AUTHOR CONTRIBUTIONS

Conceptualization: T.I., C.B., T.Pisitkun., A.J., and P.A. ; Project administration: T.I., S.B., C.B., T.Pisitkun., and S.A. ; Funding acquisition: C.B. and T.Pisitkun. ; Methodology: T.I., T.J., N.Pojdoung., N.Meesiripan., S. Sakarin., C.B., T.W., T.Phakham., T.A., C.A., P.S., P.M., M.X.T., S.Tongchusak., C.S., W.R., Q.D.L., S.D.P., K.V., S.N., K.S., R.V., N. Pesirikan, L.N., J. P., T.N., B.M., K.C., P.A., K.A., P.P., P.O., S.Sirilak., B.U., S. Sapsutthipas., S.Trisiriwanich.,T.S., A.U., N.Mingngamsup., and S.P. ; Data curation: T.I., C.B., T.W., T.A., T.Phakham., C.A., L.W., and C.S.; Data analysis: T.I., C.B., T.W., T.Phakham., T.A., C.A., T.Pisitkun., L.W., and C.S. ; Illustrations: T.W. and T. Phakham. ; Writing–review & editing manuscript: T.I., C.B., T.Phakham., C.A., B.S., T.Pisitkun., and A.J.

