## Supplemental Methods for "A randomized, placebo-controlled trial of a nasal spray solution containing broadly potent neutralizing antibodies against SARS-CoV-2 variants in healthy volunteers"

### Supplemental Methods for Biocompatibility Studies

The biocompatibility studies comprised 5 tests as described below.

1. ***In vitro* cytotoxicity study by direct contact method**

The test and control items were used as such. Sterile filter paper measuring 2.5 $cm^2$ (2.5 cm x 1 cm) was loaded with the test item without any dilution and placed at the center of each culture flask covering approximately 10% of the culture area under sterile condition, as required by the guideline ISO 10993-5:2009. Control items, sterilized high density polyethylene (HDPE) (negative control) and natural rubber latex (positive control) measuring 2.5 $cm^2$ (2.5 cm x 1 cm) were used as such and placed at the center of each culture flask. Triplicate cultures were set up for negative control, test item and positive control. This fulfills the requirement of ISO 10993, Part 5:2009 and ISO 10993-12:2012 and ISO 10993-12:2021.

Exponentially growing Balb/c 3T3 cells were seeded in culture flasks at a concentration of 1 x $10^5$ cells per mL and maintained at 5% $CO_2$ at 37 ± 1 ºC, > 90% humidity for 24 h. On the day of treatment, after confirming confluency (80%) fresh medium was replaced in each culture flask. Then, the test item, HDPE and natural rubber latex was placed at the center of each culture flask, covering approximately 10% of the culture area under sterile condition. Triplicate cultures were set up for negative control, test item and positive control. After 24 h, qualitative evaluation was performed.

Qualitative evaluation

The cultures treated with the negative control did not show any cytotoxic response (grade 0) whereas the positive control showed a severe positive cytotoxic response (grade 4). The assay was therefore considered valid. No detectable zone around or under specimen was observed in the cultures treated with test item (grade 0).

Quantitative evaluation

Viability greater than 70% was observed in the cultures treated with test item, negative control. The positive control performed as expected.

2. **Skin sensitization study in guinea pigs**

The test item was used at four different concentrations for the preliminary study (25% v/v, 50% v/v, 75% v/v in physiological saline and 100%). This fulfils the requirement of ISO 10993-12:2012 and ISO 10993-12:2021.

Initially, the range finding study (preliminary test) was conducted using four animals to determine the highest dose concentration that are well tolerated to cause only mild to moderate irritation and to determine the highest non-irritant dose of the test item. Physiological saline was used as a negative control. The four different concentrations (25% v/v, 50% v/v, 75% v/v in physiological saline and 100%) of the test item were applied (0.5 mL) as a topical patch at two different sites to all the four animals (one animal/concentration). Patches were held in contact with skin for 24 h by means of occlusive dressing. Skin reaction grading was performed using Magnusson and Kligman scale. After patch removal, no reaction or systemic effects were observed at 25% v/v, 50% v/v, 75% v/v and 100% in all the animals. Based on the results of the range finding study, 100% concentration (test item as such without any dilution) was selected for the main study (topical induction and challenge phase) and based on the injectability, 100% concentration was selected for intradermal induction.

In main study, animals were divided into two groups; G1 - five guinea pigs for control, G2 - ten guinea pigs for test item. The fur over the treatment sites were clipped and shaved on day of treatment. Induction of sensitization was a two-stage procedure with intradermal injections administered on day 0 (with FCA, vehicle and test item). No local irritation (erythema & oedema) was found at test site B (test item treated site) in the intradermal induction phase. Since no irritation was observed, pre-treatment with sodium lauryl sulphate was performed on day 6. On day 7, topical patch measuring 8 $cm^2$ loaded with 0.5 mL of test item and physiological saline was applied topically in the test and control groups respectively, on the same site as that of intradermal injections. The over patch was covered loosely with an occlusive dressing which was held in place for 48 h.

On day 21, challenge patch measuring 8 $cm^2$ loaded with 0.5 mL of test item and physiological saline was applied topically in the left and right flank of each animal for 24 h. Skin reaction grading was performed using Magnusson and Kligman scale at 24 h and 48 h, after removing the challenge patch according to ISO 10993-10:2021.

3. **Intracutaneous reactivity test in New Zealand white rabbits**

Since the test item is a liquid, it was used as such without any dilution. Physiological saline was used as negative control. This fulfils the requirement of ISO 10993-12:2012 and ISO 10993-12:2021.

About 16 h and 21 minutes prior to intracutaneous injections, fur on all the rabbits were closely clipped off their backs, allowing sufficient distance on both sides of the spine for injection. Test item and negative control were injected intracutaneously (0.2 mL of injection at five test sites/treatment). Animals were observed at 24, 48, and 72 h for morbidity, mortality and abnormal clinical signs and symptoms after injection. The skin reactions were visually scored according to ISO 10993-23:2021, at 24 h, 48 h and 72 h post injection. Observations were graded on a numerical scale for both the test item and control as shown in the following table;

| Reaction | Numerical grading |
|---|---|
| **Erythema and eschar formation** | |
| No erythema | 0 |
| Very slight erythema (barely perceptible) | 1 |
| Well-defined erythema | 2 |
| Moderate erythema | 3 |
| Severe erythema (beet-redness) to eschar formation preventing grading of erythema | 4 |
| **Oedema formation** | |
| No oedema | 0 |
| Very slight oedema (barely perceptible) | 1 |
| Well-defined oedema (edges of area well defined by definite raising) | 2 |
| Moderate oedema (raised approximately 1 mm) | 3 |
| Severe oedema (raised more than 1 mm extending beyond exposure area) | 4 |
| **Maximal possible score for irritation** | 8 |

Source: ISO 10993- Part 23: 2021

After 72 h grading, all erythema and oedema grades at 24 h, 48 h and 72 h were totalled for each test item or control for each individual animal. For calculating the score of a test item and control on each individual animal, the derived value was divided each of the totals by 15 (3 scoring periods x 5 test or control sample injection sites). To determine the overall mean score for each test item and each corresponding control, the scores for the 3 animals were added and divided by three.

The final test item score was obtained by subtracting the score of the control from the test item score.

| Negative control | Mean Reaction Score for test item | Mean Reaction Score for negative control | Overall difference (Test - Negative control) |
|---|---|---|---|
| Physiological saline | A | B | (A-B) |

If the difference between the mean reaction grades (erythema/oedema) for the test item and the control is greater than 1.0, then the test item was considered to cause intracutaneous reactivity.

**4. Acute systemic toxicity test in swiss albino mice**

Since the test item is a liquid, it was used as such without any dilution. Physiological saline was used as negative control. This fulfils the requirements of ISO 10993-12:2012 and ISO 10993-12:2021.

Two groups of mice, each comprising of five males were used for this study. Group 1 animals were treated orally using 1 mL syringe with negative control (physiological saline). Similarly, Group 2 animals were treated orally with test item (as such) without any dilution. The details are given in the following table:

| Group No. | No. of animals | Sample | Route of administration | Dose Volume |
|---|---|---|---|---|
| G1 | 5 | Physiological saline (Negative control) | Oral | 50 mL/Kg b.w. |
| G2 | 5 | Test item | Oral | 50 mL/Kg b.w. |

Animals were observed daily for mortality and morbidity throughout the experiment. Body weights of each animal were recorded prior to dosing, at 24 ± 2 h, 48 ± 2 h and 72 ± 2 h following test item administration. Clinical observation was monitored at the time of test item administration (0 h), then within 30 min and at 4 h, 24 ± 2 h, 48 ± 2 h and 72 ± 2 h following the test item administration for any clinical signs of toxicity.

**5. Subacute (28-days) systemic toxicity study in Wistar rats**

According to OECD 407, a limit test dose of 1000 mg/kg/day was selected for this study. The equivalent weight of 1 mL of the test item was determined and the volume was adjusted to obtain 1000 mg. This volume was made up to 10 mL with sterile physiological saline; animals in the test group were dosed at 10 mL/kg body weight.

All the animals were fasted overnight throughout the study period. The test item was administered to rat daily once via oral route for a period of 28 days using oral gavage as given in the table below:

| **Group No.** | **No. & Sex of animals** | **Dose** | **Route of Administration** | **Dose Volume** |
|---|---|---|---|---|
| G1 – Negative control<br>G2 - Test item | 6 M + 6 F<br>6 M + 6 F | Physiological saline<br>Test item | Oral | 10 mL/Kg b.w * |

* Administration of Substances to Laboratory Animals: Routes of Administration and Factors to Consider, J Am Assoc Lab Anim Sci. 2011; M-Male; F-Female.

Animals were observed daily for mortality, morbidity and signs/symptoms of toxicity. Body weight and feed consumption were recorded weekly. At the end of the test period (28 days), blood samples were collected for haematological and biochemical analysis. Animals were then euthanized and necropsy was performed which includes careful examination of the external surface of the body, all orifices, and the cranial, thoracic and abdominal cavities and their contents was conducted. Organs were weighed for group comparison and processed for histopathology evaluation as per Tier I listed in Annex F of ISO 10993-11:2017. Clinical observations were made at least once in a day, throughout the study period, preferably at the same time of each day and recorded. Signs noted include, but not limited to the following:

| Clinical Observation | Code No. | Observed Sign |
|---|---|---|
| Respiratory | 1 | Dyspnoea |
| | 2 | Abdominal breathing |
| | 3 | Gasping |
| | 4 | Apnoea |
| | 5 | Cyanosis |
| | 6 | Tachypnea |
| | 7 | Nostril discharges |
| Motor activities | 8 | Catatonia |
| | 9 | Somnolence |
| | 10 | Anaesthesia |
| | 11 | Catalepsy |
| | 12 | Ataxia |
| | 13 | Unusual locomotion |
| | 14 | Prostration |
| | 15 | Tremors |
| | 16 | Fasciculation |
| Convulsion | 17 | Clonic |
| | 18 | Tonic |
| | 19 | Tonic-Clonic |
| | 20 | Asphyxial |
| | 21 | Opisthotonos |
| Reflexes | 22 | Corneal |
| | 23 | Righting |
| | 24 | Myotact |
| | 25 | Light |
| | 26 | Startle reflex |
| Ocular signs | 27 | Lacrimation |
| | 28 | Miosis |
| | 29 | Mydriasis |
| | 30 | Exophthalmos |
| | 31 | Ptosis |
| | 32 | Corneal opacity |
| | 33 | Iritis |
| | 34 | Conjunctivitis |
| | 35 | Chromodacryorrhea |
| | 36 | Relaxation of nictitating membrane |
| Cardiovascular signs | 37 | Bradycardia |
| | 38 | Tachycardia |
| | 39 | Arrhythmia |
| | 40 | Vasodilation |
| | 41 | Vasoconstriction |
| Salivation | 42 | Excessive |
| Piloerection | 43 | Rough hair |
| Analgesia | 44 | Decrease reaction |
| Muscle tone | 45 | Hypotonia |
| | 46 | Hypertonia |
| Gastrointestinal | 47 | Constipation |
| | 48 | Diarrhoea |
| | 49 | Retching |
| | 50 | Emesis |
| Urinary | 51 | Hematuria |
| | 52 | Diuresis |
| Skin | 53 | Edema |
| | 54 | Erythema |

#### Hematology and Clinical Biochemistry

At the end of experiment, the blood samples were collected (after anaesthesia) prior to humanely sacrificing the animals. The haematological examination includes:

1. Hematocrit

2. Clotting potential (PT & APTT)
3. Hemoglobin concentration
4. Red blood cell count
5. White blood cell count
6. WBC differential count
7. Platelet count

Clinical biochemistry determinations to investigate major toxic effects in tissues and, specifically, effects on kidney and liver, were performed on blood samples. Determinations in plasma or serum included the following:

1. Albumin
2. ALP
3. ALT
4. AST
5. Calcium
6. Chloride
7. Cholesterol
8. Creatinine
9. GGT
10. Glucose
11. Inorganic phosphorus
12. Potassium
13. Sodium
14. Total bilirubin
15. Total protein
16. Triglycerides
17. Urea nitrogen

Urinalysis was performed during the last week of the experiment using timed (16 h to 24 h) urine volume collection. The following parameters were analysed.

1. Appearance
2. Volume
3. Sediment
4. Urobilinogen
5. Bilirubin
6. Glucose
7. Ketones
8. Occult Blood
9. Protein
10. Nitrite
11. Leukocytes
12. pH
13. Specific gravity or osmolality

Since no test item related clinical signs were observed throughout the study period, additional parameters including enzymes and immunoglobulin levels were not performed.

**Organ weight**

Liver, kidneys, adrenals, testes, epididymides, uterus, ovaries, thymus, spleen, brain (includes cerebrum, cerebellum and pons) and heart of all animals (apart from those found moribund and/or inter currently killed) were trimmed of any adherent tissue, as appropriate, and their wet weight was measured as soon as possible after dissection to avoid drying. Paired organs were weighed together. Relative weight of individual organ was calculated by dividing the absolute organ weight with the body weight of the animal and multiplying by 100. Organ weight of animals that are found dead during the study was not recorded.

**Histopathology**

The following list of organs/tissues (Tier II) were collected and preserved from all the animals as mentioned in Annex E of ISO 10993-11:2017.

1. Adrenals
2. All gross lesions (including treatment sites)
3. Aorta
4. Bone marrow (sternum)
5. Brain (including cerebrum, cerebellum and pons)
6. Caecum
7. Colon
8. Duodenum
9. Epididymis
10. Oesophagus
11. Eyes
12. Femur
13. Heart
14. Ileum
15. Jejunum
16. Kidneys
17. Liver
18. Lungs & bronchi
19. Lymph nodes (cervical and mesenteric)
20. Mammary gland (female)
21. Muscle (skeletal)
22. Nerve (sciatic)
23. Ovaries
24. Pancreas
25. Parathyroid
26. Pituitary
27. Prostate
28. Rectum
29. Salivary glands
30. Seminal vesicles
31. Skin
32. Spinal cord
33. Spleen
34. Stomach
35. Testes
36. Thymus
37. Thyroid
38. Trachea
39. Urinary Bladder
40. Uterus
41. Vagina

Limited histopathological analysis was conducted for below mentioned organs/tissues (Tier I) in the control and treated groups as listed in Annex F of ISO 10993-11:2017.

1. Heart
2. Liver
3. Adrenals
4. Kidneys

5. Skin
6. Spleen
7. Muscle
8. Brain
9. Testes/ Ovaries
10. Lungs and Bronchi
11. Femur
12. Bone marrow (sternum)
