## Supplemental Data S1 for "A randomized, placebo-controlled trial of a nasal spray solution containing broadly potent neutralizing antibodies against SARS-CoV-2 variants in healthy volunteers"

### Supplemental Data S1
### Biocompatibility results

#### 1. *In vitro* cytotoxicity using the direct contact method

Before treatment, approximately 80% confluency was confirmed in all the cultures. In the qualitative evaluation, the cells treated with the positive control showed a complete destruction of cell layers (grade 4). No such destruction was evident in the negative control. Quantitatively, the cells treated with the negative control had a mean viability of 100% and positive control had 13.48% viability. Based on qualitative and quantitative evaluations, the assay was considered as valid.

- **Qualitative evaluation**

The cultures treated with the negative control did not show any cytotoxic response (grade 0) whereas the positive control showed a severe positive cytotoxic response (grade 4). The assay was therefore considered valid. The cultures treated with test item showed no detectable zone around or under specimen which was considered non-cytotoxic (grade 0) Table 1.

**Table 1 Qualitative evaluation**

| Sample | Culture | Reactivity | Grade |
|---|---|---|---|
| Negative Control | 1 | None | 0 |
| | 2 | None | 0 |
| | 3 | None | 0 |
| Test item | 1 | None | 0 |
| | 2 | None | 0 |
| | 3 | None | 0 |
| Positive control | 1 | Severe | 4 |
| | 2 | Severe | 4 |
| | 3 | Severe | 4 |

- **Quantitative evaluation**

Viability greater than 70% was observed in the cultures treated with test item, negative control. The positive control performed as expected (Table 2).

**Table 2 Quantitative evaluation (NR absorbance at 550 nm)**

| Sample | Replicate 1 | Replicate 2 | Replicate 3 | Mean | Viability (%) | Cytotoxicity (%) |
|---|---|---|---|---|---|---|
| Negative control | 0.88 | 0.90 | 0.90 | 0.89 | 100.00 | 0.00 |
| Test item | 0.86 | 0.85 | 0.81 | 0.84 | 94.38 | 5.62 |
| Positive control | 0.14 | 0.10 | 0.11 | 0.12 | 13.48 | 86.52 |

#### 2. Skin sensitization using the guinea pig maximization test

- **Mortality & morbidity**

No mortality or morbidity occurred in any of the animals used in this study.

Body weight A gradual increase in the body weight was observed in all the animals at the end of the experiment. Body weight of animals recorded prior to dosing and end of the experiment are given in Table 3.

**Table 3 Individual body weight of all animals in skin sensitization test**

| Test/Group | Sex | Animal No. | Weight (in grams) | | |
|---|---|---|---|---|---|
| | | | At the time of dosing | At the end of experiment | Increase in weight |
| Preliminary Test | M | 01 | 376.34 | 378.36 | 2.02 |
| | | 02 | 345.09 | 347.00 | 1.91 |
| | | 03 | 358.41 | 360.57 | 2.16 |
| | | 04 | 345.19 | 347.16 | 1.97 |
| | | Mean ± SD | 356.26 ± 14.78 | 358.27 ± 14.83 | 2.02 ± 0.11 |
| Main Test G1 | M | 01 | 341.25 | 373.76 | 32.51 |
| | | 02 | 368.37 | 400.29 | 31.92 |
| | | 03 | 376.13 | 409.15 | 33.02 |
| | | 04 | 348.29 | 379.99 | 31.70 |
| | | 05 | 365.39 | 394.17 | 28.78 |
| | | Mean ± SD | 359.89 ± 14.56 | 391.47 ± 14.52 | 31.59 ± 1.65 |
| Main Test G2 | M | 06 | 346.24 | 379.10 | 32.86 |
| | | 07 | 341.37 | 374.38 | 33.01 |
| | | 08 | 368.92 | 398.32 | 29.40 |
| | | 09 | 357.80 | 386.60 | 28.80 |
| | | 10 | 376.60 | 406.69 | 30.09 |
| | | 11 | 351.63 | 383.41 | 31.78 |
| | | 12 | 348.87 | 380.45 | 31.58 |
| | | 13 | 361.42 | 393.71 | 32.29 |
| | | 14 | 339.02 | 368.13 | 29.11 |
| | | 15 | 372.27 | 402.35 | 30.08 |
| | | Mean ± SD | 356.41 ± 13.14 | 387.31 ± 12.60 | 30.90 ± 1.59 |

M - Male; SD- Standard Deviation

- **Grading of skin reactions**

Grading of skin reactions performed at 24 h and 48 h after removing the challenge patch are given in Table 4. No sensitization reactions were observed in animals treated with the negative control. No evidence of sensitization was seen in any of the test item treated animals, as no skin reactions were observed.

**Table 4 Grading of skin reaction**

| Group | Sex | Animal No. | After 24 h exposure |
|---|---|---|---|
| Preliminary Test | M | 01 | 0 |
| | | 02 | 0 |
| | | 03 | 0 |
| | | 04 | 0 |

| Group | Sex | Animal No. | Magnusson and Kligman Scale | | | |
|---|---|---|---|---|---|---|
| | | | 24 h | | 48 h | |
| | | | C | T | C | T |
| Main Test G1 | M | 01 | 0 | 0 | 0 | 0 |
| | | 02 | 0 | 0 | 0 | 0 |
| | | 03 | 0 | 0 | 0 | 0 |
| | | 04 | 0 | 0 | 0 | 0 |
| | | 05 | 0 | 0 | 0 | 0 |
| Main Test G2 | M | 06 | 0 | 0 | 0 | 0 |
| | | 07 | 0 | 0 | 0 | 0 |
| | | 08 | 0 | 0 | 0 | 0 |
| | | 09 | 0 | 0 | 0 | 0 |
| | | 10 | 0 | 0 | 0 | 0 |
| | | 11 | 0 | 0 | 0 | 0 |
| | | 12 | 0 | 0 | 0 | 0 |
| | | 13 | 0 | 0 | 0 | 0 |
| | | 14 | 0 | 0 | 0 | 0 |
| | | 15 | 0 | 0 | 0 | 0 |

M-Male; C- Control site; T- Treated site; h- hour

3. **Intracutaneous reactivity potentials in New Zealand white rabbits**

- **Mortality & Morbidity**

No mortality or morbidity occurred in any of the animals throughout the experiment.

- **Body Weight**

A gradual increase in body weight was observed in all the animals at the end of experiment. Individual body weight of the animals is given in Table 5.

**Table 5 Individual body weights of New Zealand white rabbits**

| Animal No. | Sex | Body weight (g) | |
|---|---|---|---|
| | | Initial | Final |
| 1 | M | 2204.0 | 2238.1 |
| 2 | | 2390.4 | 2427.9 |
| 3 | | 2162.1 | 2190.6 |

M-Male

- **Clinical Observation**

No signs of ill health or overt toxicity were observed in any of the test animals.

- **Scoring of Skin Reaction**

Test item injected sites showed erythema at 24 h, 48 h and 72 h and no reactions were observed at the control sites. Grading of skin reactions for individual animals were given in Table 6. The difference of the mean skin reaction scores for the test item and control was 0.4 (Table 7). The overall difference between the mean reaction grades (erythema) for the test item and the control is less than 1.0.

**Table 6 Grading of skin reactions for individual New Zealand White rabbits**

| Time points | | | 24 h | | | | 48 h | | | | 72 h | | | |
|---|---|---|---|---|---|---|---|---|---|---|---|---|---|---|
| **Animal No.** | **Sex** | **Sites** | **Test item** | | **Negative Control** | | **Test item** | | **Negative Control** | | **Test item** | | **Negative Control** | |
| | | | **E** | **O** | **E** | **O** | **E** | **O** | **E** | **O** | **E** | **O** | **E** | **O** |
| **1** | **Male** | 1 | 1 | 0 | 0 | 0 | 1 | 0 | 0 | 0 | 0 | 0 | 0 | 0 |
| | | 2 | 0 | 0 | 0 | 0 | 1 | 0 | 0 | 0 | 0 | 0 | 0 | 0 |
| | | 3 | 0 | 0 | 0 | 0 | 0 | 0 | 0 | 0 | 0 | 0 | 0 | 0 |
| | | 4 | 1 | 0 | 0 | 0 | 0 | 0 | 0 | 0 | 0 | 0 | 0 | 0 |
| | | 5 | 1 | 0 | 0 | 0 | 1 | 0 | 0 | 0 | 0 | 0 | 0 | 0 |
| | | E+O | 3 | | 0 | | 3 | | 0 | | 0 | | 0 | |
| | | E+O/ 5 sites | 0.6 | | 0 | | 0.6 | | 0 | | 0 | | 0 | |
| **2** | | 1 | 0 | 0 | 0 | 0 | 1 | 0 | 0 | 0 | 0 | 0 | 0 | 0 |
| | | 2 | 1 | 0 | 0 | 0 | 1 | 0 | 0 | 0 | 0 | 0 | 0 | 0 |
| | | 3 | 1 | 0 | 0 | 0 | 0 | 0 | 0 | 0 | 0 | 0 | 0 | 0 |
| | | 4 | 0 | 0 | 0 | 0 | 0 | 0 | 0 | 0 | 1 | 0 | 0 | 0 |
| | | 5 | 0 | 0 | 0 | 0 | 0 | 0 | 0 | 0 | 1 | 0 | 0 | 0 |
| | | E+O | 2 | | 0 | | 2 | | 0 | | 2 | | 0 | |
| | | E+O/ 5 sites | 0.4 | | 0 | | 0.4 | | 0 | | 0.4 | | 0 | |
| **3** | | 1 | 1 | 0 | 0 | 0 | 0 | 0 | 0 | 0 | 0 | 0 | 0 | 0 |
| | | 2 | 1 | 0 | 0 | 0 | 0 | 0 | 0 | 0 | 0 | 0 | 0 | 0 |
| | | 3 | 0 | 0 | 0 | 0 | 0 | 0 | 0 | 0 | 0 | 0 | 0 | 0 |
| | | 4 | 0 | 0 | 0 | 0 | 1 | 0 | 0 | 0 | 0 | 0 | 0 | 0 |
| | | 5 | 1 | 0 | 0 | 0 | 0 | 0 | 0 | 0 | 0 | 0 | 0 | 0 |
| | | E+O | 3 | | 0 | | 1 | | 0 | | 0 | | 0 | |
| | | E+O/ 5 sites | 0.6 | | 0 | | 0.2 | | 0 | | 0 | | 0 | |

E, Erythema; O, Oedema

**Table 7 Calculation of skin reactions - Overall mean score and difference**

| Negative control | Test item | Negative control | Overall difference |
|---|---|---|---|
| | E+O | E+O | Test item - negative control |
| Physiological saline | 0.4 | 0 | 0.4 |

E, Erythema; O, Oedema

4. **Acute systemic toxicity study via intranasal in mice**
   - **Mortality & Morbidity**

No mortality or morbidity were observed in any of the animals used in this study.

- **Body Weight**

A gradual increase in body weight was observed in all animals at the end of experiment. Individual body weight of the animals is given in Table 8.

**Table 8 Individual body weights**

| Group No. | Animal No. | Sex | Weight (in grams) | | | |
|---|---|---|---|---|---|---|
| | | | Days | | | |
| | | | Day 0 | 24 h | 48 h | 72 h |
| G1 | 1 | M | 17.88 | 18.08 | 18.24 | 18.40 |
| | 2 | | 18.45 | 18.57 | 18.77 | 18.94 |
| | 3 | | 18.76 | 18.88 | 18.99 | 19.19 |
| | 4 | | 20.57 | 20.75 | 20.89 | 21.06 |
| | 5 | | 21.26 | 21.43 | 21.61 | 21.72 |
| G2 | 6 | M | 18.07 | 18.19 | 18.33 | 18.43 |
| | 7 | | 18.66 | 18.79 | 18.94 | 19.13 |
| | 8 | | 19.40 | 19.54 | 19.70 | 19.87 |
| | 9 | | 20.95 | 21.07 | 21.23 | 21.36 |
| | 10 | | 21.29 | 21.47 | 21.57 | 21.67 |

M - Male; G - Group

- **Clinical Observation**

No signs of ill health or overt toxicity were observed in any of the animals (Table 9).

**Table 9 Clinical Observations**

| Group No. | Animal No. | Sex | Observation at | | | | | |
|---|---|---|---|---|---|---|---|---|
| | | | 0 h | 30 min | 4 h | 24 h | 48 h | 72 h |
| G1 | 1 | M | N | N | N | N | N | N |
| | 2 | | N | N | N | N | N | N |
| | 3 | | N | N | N | N | N | N |
| | 4 | | N | N | N | N | N | N |
| | 5 | | N | N | N | N | N | N |
| G2 | 6 | M | N | N | N | N | N | N |
| | 7 | | N | N | N | N | N | N |
| | 8 | | N | N | N | N | N | N |
| | 9 | | N | N | N | N | N | N |
| | 10 | | N | N | N | N | N | N |

M - Male; G - Group; h-hour; min- minutes; N-Normal

- **Gross Pathology, Clinical Pathology, and Histopathology**

Since no abnormal clinical signs were observed, no gross pathology, clinical pathology or histopathology were conducted.

5. **28-day subacute systemic toxicity study via oral administration in rats**

- **Mortality & Morbidity**

No morbidity or mortality occurred in any of the animals in both the control and treated group throughout the study.

- **Body Weight**

A gradual increase in body weight was observed in all the animals and no significant changes were observed between the treated and control groups as shown in Table 10.

**Table 10 Summary of weekly body weight (g)**

| Group | Days | | | | |
|---|---|---|---|---|---|
| | 0 | 7 | 14 | 21 | 28 |
| G1 Male N = 06 | 186.07 ± 8.502 | 205.73 ± 9.330 | 225.08 ± 8.465 | 244.25 ± 9.273 | 263.61 ± 8.898 |
| G1 Female N = 06 | 181.36 ± 6.732 | 202.14 ± 8.191 | 222.34 ± 7.872 | 242.06 ± 8.344 | 262.41 ± 9.658 |
| G2 Male N = 06 | 187.36 ± 7.350 | 207.94 ± 6.504 | 228.65 ± 5.167 | 250.61 ± 6.977 | 270.99 ± 6.138 |
| G2 Female N = 06 | 182.38 ± 6.877 | 201.12 ± 7.429 | 220.21 ± 7.577 | 238.98 ± 9.434 | 257.12 ± 9.863 |

Values are expressed as Mean ± SD; N, Number of animals.

- **Feed consumption**

None of the animals in treated group showed significant difference in the feed consumption when compared to the control group as shown in Table 11.

**Table 11 Summary of weekly feed consumption (g)**

| Group/Sex | Days | | | |
|---|---|---|---|---|
| | 7 | 14 | 21 | 28 |
| G1 Male N = 06 | 94.70 ± 3.737 | 99.45 ± 6.117 | 97.86 ± 3.388 | 98.77 ± 4.728 |
| G1 Female N = 06 | 96.73 ± 3.096 | 98.90 ± 5.077 | 100.51 ± 4.144 | 98.04 ± 3.906 |
| G2 Male N = 06 | 95.95 ± 5.377 | 99.26 ± 3.163 | 95.36 ± 3.615 | 98.39 ± 5.488 |
| G2 Female N = 06 | 98.77 ± 4.728 | 97.38 ± 4.902 | 99.19 ± 5.394 | 97.75 ± 6.796 |

Values are expressed as Mean ± SD

- **Clinical Observation**

None of the animals from any group showed any clinical signs of toxicity during the course of the study.

- **Urinalysis**

No abnormal difference was observed in the urine analysis performed in the samples and all the parameters were within acceptable range when compared to the control animals see in Table 12.

- **Hematology and Clinical Biochemistry**

In male animals, no significant changes were observed in haematology, biochemical parameters and electrolytes when compared with the control group.

In female animals, no significant changes were observed in haematology (except WBC values), biochemical parameters and electrolytes. A significant difference was observed in the WBC values (p=0.0027) between the control and treated groups. Since the values were within the normal physiological range, these changes were considered not related to the test item as shown in Table 13.

- **Gross Pathology**

No test item and control item related gross pathological changes were observed. Most tissues were macroscopically unremarkable, and the other findings seen were generally consistent with the usual pattern of findings in animals of this strain and age as shown in Table 14.

- **Organ Weights**

In male animals, no significant changes were observed in absolute and relative organ weights (except relative organ weights of adrenal and epididymis) when compared with the control group. A significant difference was observed in the relative organ weight of adrenal (p=0.0269) and epididymis (p=0.0277) between the control and treated groups. Since the values were within the normal physiological range, these changes were considered not related to the test item as shown in Table 15.

- **Histopathology**

No test item and control item related histopathological changes were observed. Other observed microscopic findings were generally infrequent, of a minor nature and consistent with the usual pattern of findings in animals of this strain and age. All of the observed changes were considered to be spontaneous background lesions and not related to adverse effects of control or test item as shown in Table 16.

Since no abnormal findings were observed in Tier I organs/tissues and in clinical pathology analysis (clinical chemistry and haematology), the histopathology analysis of Tier II organs/tissues were not performed.

**Table 12 Summary of Urinalysis**

| ORGANS | Parameter | GROUPS | | | |
|---|---|---|---|---|---|
| | | G1 | G1 | G2 | G2 |
| | | Male | Female | Male | Female |
| | | 6 animals | 6 animals | 6 animals | 6 animals |
| Volume | Mean | 6.97 | 6.65 | 6.28 | 7.30 |
| | SD | 0.871 | 0.536 | 0.546 | 1.126 |
| Colour | Pale Yellow | 3 | 2 | 3 | 3 |
| | Yellow | 3 | 3 | 2 | 2 |
| | Brown | 0 | 1 | 1 | 1 |
| Urobilinogen | 3.3 | 4 | 4 | 3 | 3 |
| | 16 | 2 | 1 | 2 | 1 |
| | 33 | 0 | 1 | 1 | 2 |
| Bilirubin | Positive | 0 | 0 | 0 | 0 |
| | Negative | 6 | 6 | 6 | 6 |
| Ketone | Negative | 6 | 6 | 6 | 6 |
| | 0.5 | 0 | 0 | 0 | 0 |
| Blood cells | Negative | 6 | 6 | 6 | 6 |
| Protein | Negative | 1 | 1 | 1 | 1 |
| | 0.2 | 2 | 4 | 4 | 3 |
| | 0.3 | 3 | 1 | 1 | 2 |
| Nitrite | Negative | 6 | 6 | 6 | 6 |
| Leucocytes | Negative | 6 | 6 | 6 | 6 |
| Glucose | Negative | 6 | 6 | 6 | 6 |
| Gravity | 1.010 | 2 | 1 | 2 | 2 |
| | 1.015 | 0 | 3 | 2 | 1 |
| | 1.020 | 3 | 1 | 1 | 1 |
| | 1.025 | 1 | 1 | 1 | 2 |
| | ≥1.030 | 0 | 0 | 0 | 0 |
| pH | 6.0 | 1 | 0 | 1 | 0 |
| | 6.5 | 3 | 2 | 2 | 1 |
| | 7.0 | 2 | 2 | 2 | 2 |
| | 7.5 | 0 | 1 | 1 | 1 |
| | 8.0 | 0 | 1 | 0 | 1 |
| | 8.5 | 0 | 0 | 0 | 1 |
| Microscopic Examination | | NAD | NAD | NAD | NAD |

**Table 13 Summary of hematology measurements**

| Group | RBC ($10^6/\mu L$) | Hb (g/dL) | HCT (%) | WBC ($10^3/\mu L$) | PLT ($10^3/\mu L$) | Differential count L (%) | Differential count M (%) | Differential count G (%) | MCH (pg) | MCHC (g/dL) | MCV (fL) |
|---|---|---|---|---|---|---|---|---|---|---|---|
| G1 Male N = 06 | 8.08 ± 0.423 | 13.95 ± 0.742 | 41.15 ± 2.015 | 12.42 ± 1.359 | 676.17 ± 90.19 | 78.05 ± 7.302 | 3.92 ± 0.240 | 18.03 ± 7.278 | 17.32 ± 1.338 | 33.97 ± 2.574 | 50.97 ± 2.023 |
| G1 Female N = 06 | 7.63 ± 0.467 | 13.62 ± 0.741 | 42.17 ± 1.468 | 13.63 ± 1.063 | 646.50 ± 65.06 | 72.27 ± 6.178 | 2.80 ± 0.684 | 24.93 ± 6.550 | 17.90 ± 1.393 | 32.33 ± 2.023 | 55.40 ± 3.125 |
| G2 Male N = 06 | 7.69 ± 0.699 | 14.25 ± 0.946 | 39.50 ± 3.010 | 12.35 ± 1.677 | 632.33 ± 74.93 | 76.47 ± 4.219 | 3.32 ± 0.898 | 20.22 ± 4.770 | 18.68 ± 2.319 | 36.30 ± 4.077 | 51.62 ± 4.573 |
| G2 Female N = 06 | 7.39 ± 0.648 | 13.07 ± 1.236 | 41.92 ± 2.949 | 11.33 ± 0.94 | 706.17 ± 43.64 | 74.70 ± 5.519 | 3.43 ± 1.042 | 21.87 ± 6.353 | 17.85 ± 2.935 | 31.27 ± 3.204 | 57.05 ± 6.344 |

Values are expressed as Mean ± SD; Hb, hemoglobin; RBC, red blood cell count; HCT, hematocrit; MCH, mean corpuscular haemoglobin; MCHC, mean corpuscular haemoglobin concentration; MCV, mean corpuscular volume; WBC, white blood cell count; L, lymphocyte percentage; M, monocyte percentage; G, granulocyte percentage; PLT, platelet count; CT, clotting time.

**Table 14 Summary of gross pathology**

| ORGANS | LESIONS | Groups | | | |
|---|---|---|---|---|---|
| | | G1 | G1 | G2 | G2 |
| | | Male | Female | Male | Female |
| | | 6 animals | 6 animals | 6 animals | 6 animals |
| **Adrenals** | NAD | - | - | - | - |
| **Aorta** | NAD | - | - | - | - |
| **Bone marrow (Sternum)** | NAD | - | - | - | - |
| **Brain** | NAD | - | - | - | - |
| **Caecum** | NAD | - | - | - | - |
| **Colon** | NAD | - | - | - | - |
| **Duodenum** | NAD | - | - | - | - |
| **Epididymis** | NAD | - | NA | - | NA |
| **Oesophagus** | NAD | - | - | - | - |
| **Eyes** | NAD | - | - | - | - |
| **Femur** | NAD | - | - | - | - |
| **Heart** | NAD | - | - | - | - |
| **Ileum** | NAD | - | - | - | - |
| **Jejunum** | NAD | - | - | - | - |
| **Kidneys** | Irregular areas of red discoloration – diffuse | - | - | 1 | - |
| **Liver** | Irregular areas of red discoloration – diffuse | - | 1 | - | 1 |
| **Lungs and Bronchi** | Irregular areas of red discoloration – diffuse | 1 | 1 | 1 | - |
| **Lymph nodes** | NAD | - | - | - | - |
| **Mammary gland** | NAD | NA | - | NA | - |
| **Skeletal muscle** | NAD | - | - | - | - |
| **Sciatic nerve** | NAD | - | - | - | - |
| **Ovaries** | NAD | NA | - | NA | - |
| **Pancreas** | NAD | - | - | - | - |
| **Parathyroid** | NAD | - | - | - | - |
| **Pituitary** | NAD | - | - | - | - |
| **Prostate** | NAD | - | NA | - | NA |
| **Rectum** | NAD | - | - | - | - |
| **Salivary glands** | NAD | - | - | - | - |
| **Seminal vesicles** | NAD | - | NA | - | NA |
| **Skin** | NAD | - | - | - | - |

| ORGANS | LESIONS | Groups | | | |
|---|---|---|---|---|---|
| | | G1 | G1 | G2 | G2 |
| | | Male | Female | Male | Female |
| | | 6 animals | 6 animals | 6 animals | 6 animals |
| **Spinal cord** | NAD | - | - | - | - |
| **Femur** | NAD | - | - | - | - |
| **Spleen** | NAD | - | - | - | - |
| **Stomach** | NAD | - | - | - | - |
| **Testes** | NAD | - | NA | - | NA |
| **Thymus** | NAD | - | - | - | - |
| **Thyroid** | NAD | - | - | - | - |
| **Trachea** | NAD | - | - | - | - |
| **Urinary bladder** | NAD | - | - | - | - |
| **Uterus** | NAD | NA | - | NA | - |
| **Vagina** | NAD | NA | - | NA | - |

NAD – No Abnormality Detected, NA – Not Applicable.

#### Table 15 Summary of relative organ weights (%)

| Group | Adrenal | Thymus | Spleen | Heart | Brain | Kidney | Liver | Testes* / Ovary# | Epididymis* / Uterus# |
|---|---|---|---|---|---|---|---|---|---|
| **G1 Male N = 06** | 0.03 ± 0.004 | 0.13 ± 0.032 | 0.29 ± 0.070 | 0.38 ± 0.119 | 0.68 ± 0.044 | 0.64 ± 0.061 | 3.79 ± 0.464 | 0.49 ± 0.107 | 0.39 ± 0.016 |
| **G1 Female N = 06** | 0.02 ± 0.005 | 0.13 ± 0.017 | 0.34 ± 0.065 | 0.33 ± 0.063 | 0.68 ± 0.068 | 0.66 ± 0.067 | 3.66 ± 0.538 | 0.059 ± 0.012 | 0.31 ± 0.036 |
| **G2 Male N = 06** | 0.02 ± 0.004 | 0.11 ± 0.039 | 0.35 ± 0.093 | 0.35 ± 0.118 | 0.64 ± 0.036 | 0.64 ± 0.020 | 3.63 ± 0.682 | 0.50 ± 0.118 | 0.37 ± 0.018 |
| **G2 Female N = 06** | 0.02 ± 0.005 | 0.12 ± 0.029 | 0.34 ± 0.076 | 0.35 ± 0.079 | 0.70 ± 0.049 | 0.72 ± 0.058 | 3.56 ± 0.223 | 0.06 ± 0.005 | 0.32 ± 0.073 |

Values are expressed in grams as Mean ± SD; * for males; # for females.

**Table 16 Summary of histopathology**

| ORGANS | LESIONS | GROUPS | | | |
|---|---|---|---|---|---|
| | | G1 | G1 | G2 | G2 |
| | | Male | Female | Male | Female |
| | | 6 animals | 6 animals | 6 animals | 6 animals |
| **Heart** | NSML | - | - | - | - |
| **Liver** | Infiltrate, inflammatory cells, focal, minimal | 1 | 1 | - | - |
| | Pigments, brown to black, diffuse, minimal | - | - | 1 | - |
| | Congestion, sinusoidal, diffuse, minimal | 1 | 2 | 1 | 1 |
| **Adrenals** | Congestion, diffuse, minimal | 1 | - | 0 | - |
| | Vacuolation, cytoplasmic, diffuse, cortex, Zona fasciculata, mild | - | - | - | 1 |
| **Kidneys** | Congestion, diffuse, minimal | - | 1 | 1 | - |
| | Infiltrate, mononuclear cells, interstitial, multifocal, cortex, mild | 1 | - | 1 | 1 |
| **Skin** | NSML | - | - | - | - |
| **Spleen** | Pigments, brown to black, multifocal, red pulp, minimal | - | 1 | 1 | 1 |
| **Skeletal muscle** | NSML | - | - | - | - |
| **Brain** | NSML | - | - | - | - |
| **Testes** | NSML | - | NA | - | NA |
| **Ovaries** | NSML | NA | - | NA | - |
| **Lungs and Bronchi** | Infiltrate, mononuclear cells, perivascular, multifocal, minimal | - | 1 | 1 | 0 |
| | Congestion, diffuse, minimal | 1 | 1 | 2 | - |
| | Congestion and oedema, diffuse, minimal | - | 1 | 1 | - |
| | Infiltrate, mononuclear cells, perivascular, multifocal, mild<br>Congestion and oedema, diffuse, minimal | 1 | - | 1 | 1 |
| **Femur** | NSML | - | - | - | - |
| **Bone marrow (sternum)** | NSML | - | - | - | - |

NSML – No significant microscopic lesions; NA – Not applicable.
