## Supplemental Data S2 for "A randomized, placebo-controlled trial of a nasal spray solution containing broadly potent neutralizing antibodies against SARS-CoV-2 variants in healthy volunteers"

### Supplemental Data S2
#### Nasal sinuscopy images of participants in the COVITRAP™ and placebo groups on days 0, 7, and 14

**Nasal sinuscopy images are displayed in random order of participants.**

#### COVITRAP™ group (n = 27)

| | Nostril | | | Nostril | |
|---|---|---|---|---|---|
| C1 | Right | Left | C2 | Right | Left |
| Day 0 | 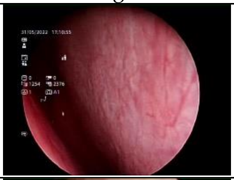 | 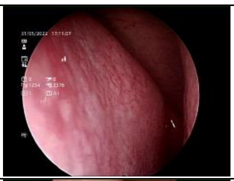 | Day 0 | 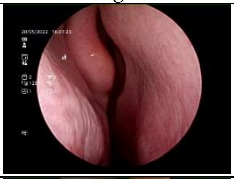 | 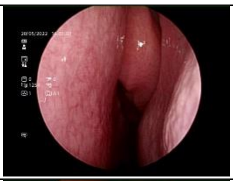 |
| Day 7 | 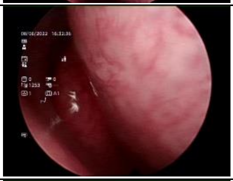 | 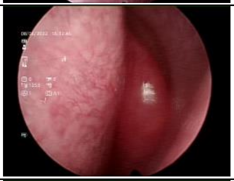 | Day 7 | 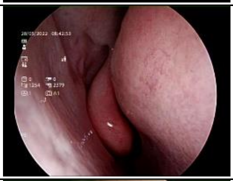 | 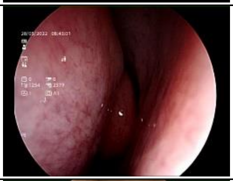 |
| Day 14 | 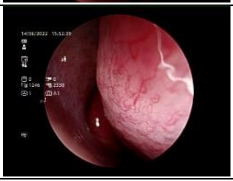 | 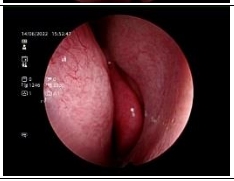 | Day 14 | 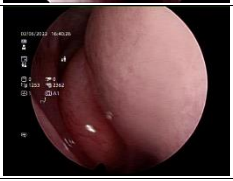 | 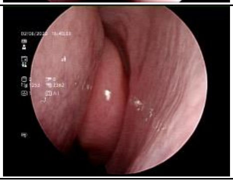 |
| C3 | Right | Left | C4 | Right | Left |
| Day 0 | 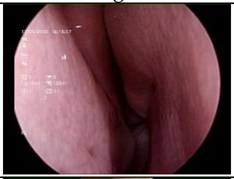 | 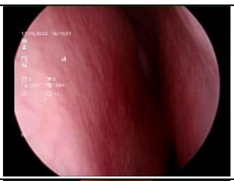 | Day 0 | 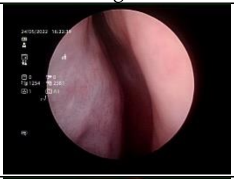 | 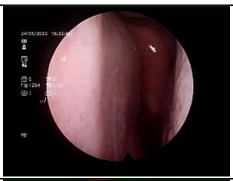 |
| Day 7 | 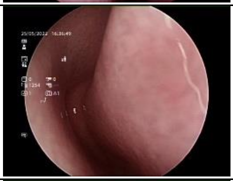 | 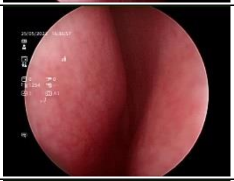 | Day 7 | 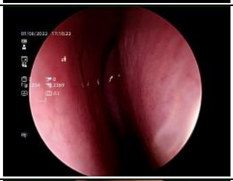 | 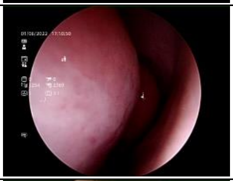 |
| Day 14 | 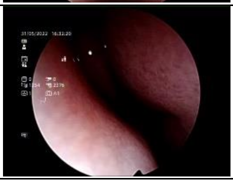 | 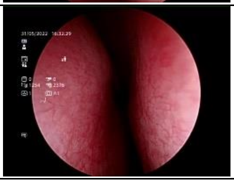 | Day 14 | 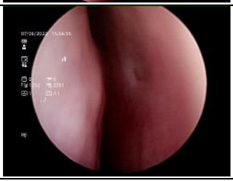 | 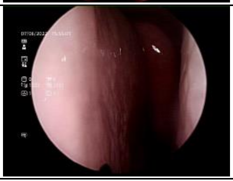 |
| C5 | Right | Left | C6 | Right | Left |
| Day 0 | 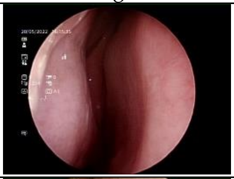 | 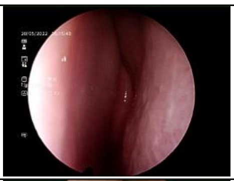 | Day 0 | 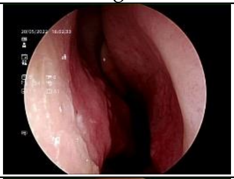 | 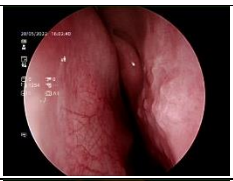 |
| Day 7 | 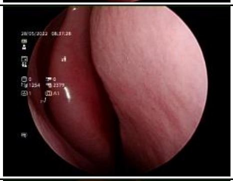 | 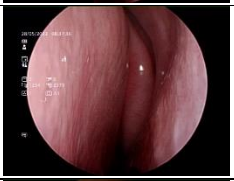 | Day 7 | 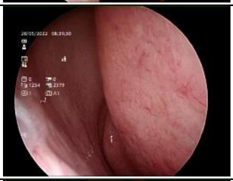 | 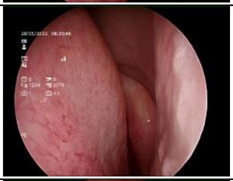 |
| Day 14 | 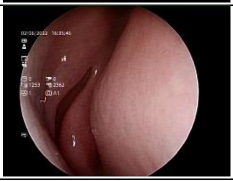 | 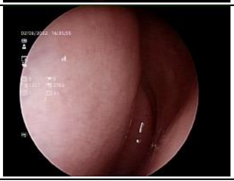 | Day 14 | 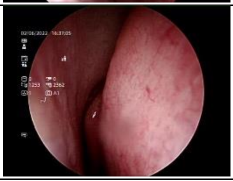 | 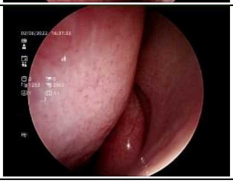 |

| | Nostril | | | Nostril | |
|---|---|---|---|---|---|
| C7 | Right | Left | C8 | Right | Left |
| Day 0 | 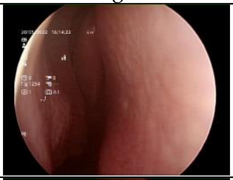 | 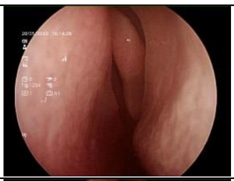 | Day 0 | 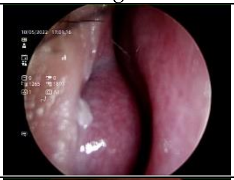 | 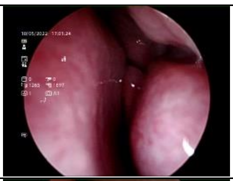 |
| Day 7 | 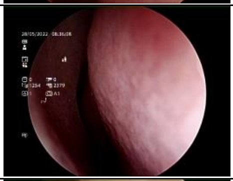 | 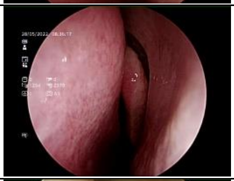 | Day 7 | 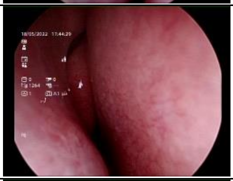 | 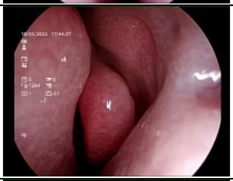 |
| Day 14 | 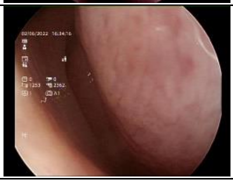 | 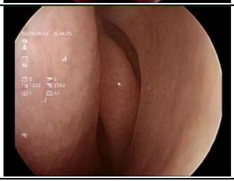 | Day 14 | 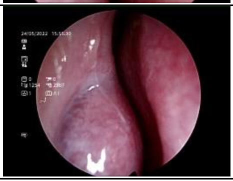 | 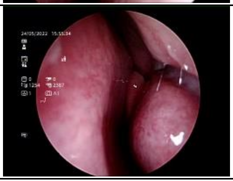 |
| C9 | Right | Left | C10 | Right | Left |
| Day 0 | 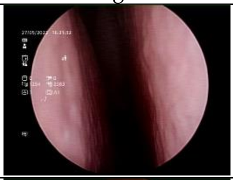 | 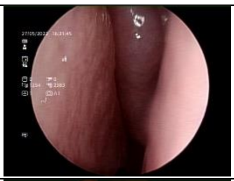 | Day 0 | 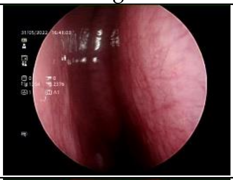 | 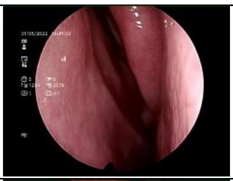 |
| Day 7 | 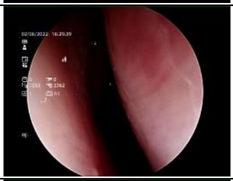 | 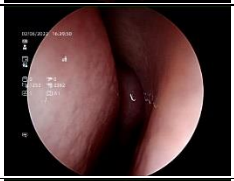 | Day 7 | 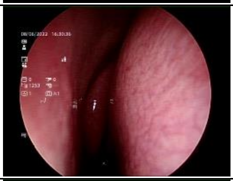 | 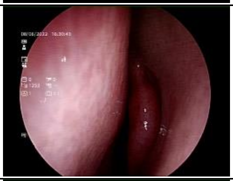 |
| Day 14 | 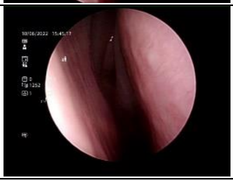 | 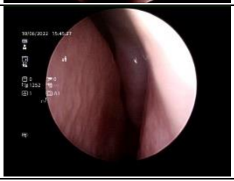 | Day 14 | 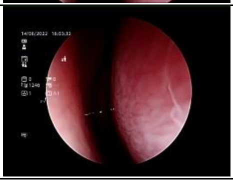 | 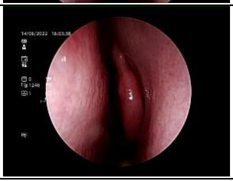 |
| C11 | Right | Left | C12 | Right | Left |
| Day 0 | 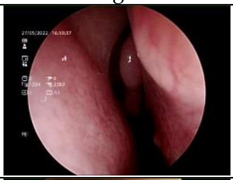 | 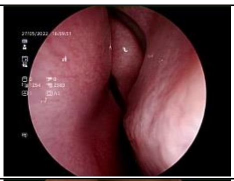 | Day 0 | 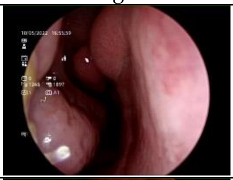 | 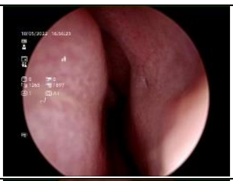 |
| Day 7 | 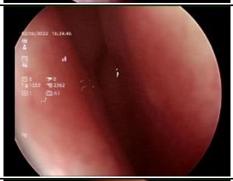 | 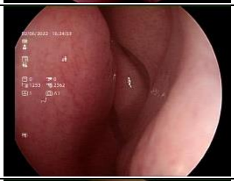 | Day 7 | 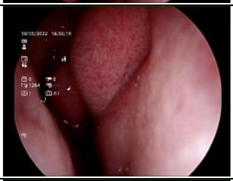 | 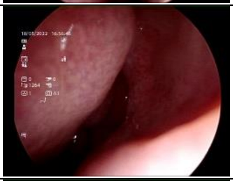 |
| Day 14 | 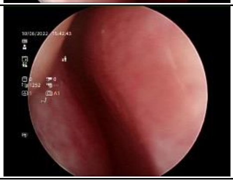 | 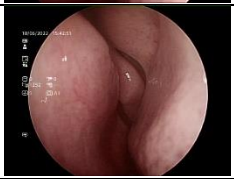 | Day 14 | 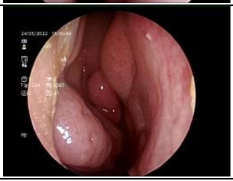 | 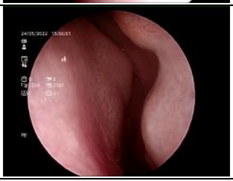 |

| | Nostril | | | Nostril | |
|---|---|---|---|---|---|
| C13 | Right | Left | C14 | Right | Left |
| Day 0 | 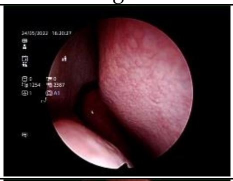 | 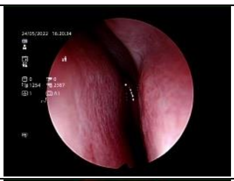 | Day 0 | 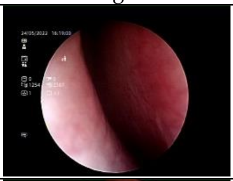 | 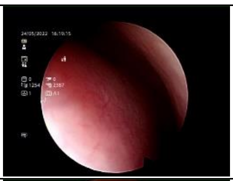 |
| Day 7 | 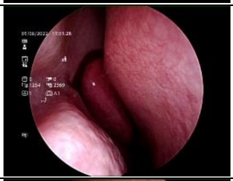 | 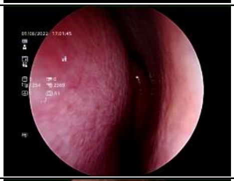 | Day 7 | 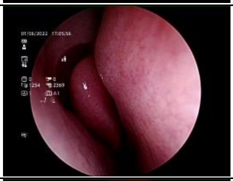 | 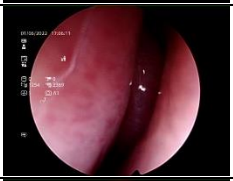 |
| Day 14 | 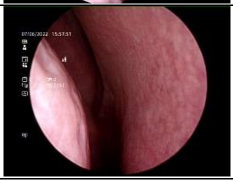 | 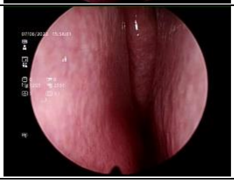 | Day 14 | 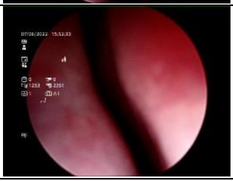 | 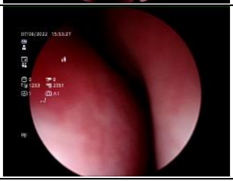 |
| C15 | Right | Left | C16 | Right | Left |
| Day 0 | 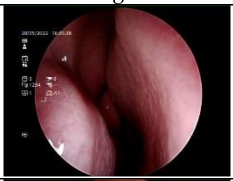 | 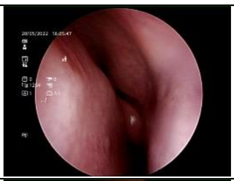 | Day 0 | 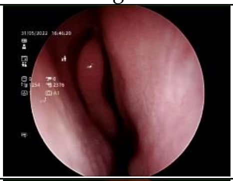 | 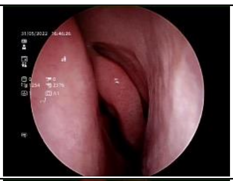 |
| Day 7 | 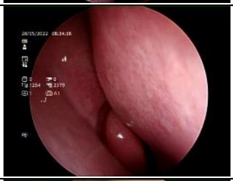 | 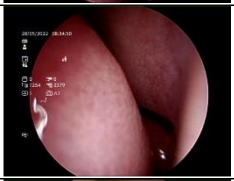 | Day 7 | 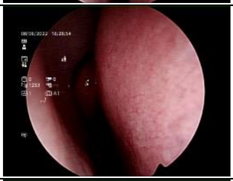 | 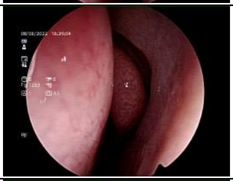 |
| Day 14 | 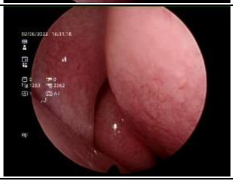 | 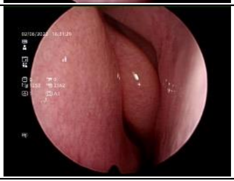 | Day 14 | 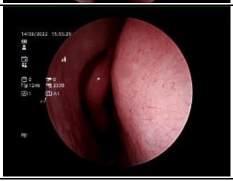 | 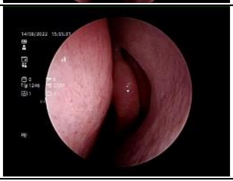 |
| C17 | Right | Left | C18 | Right | Left |
| Day 0 | 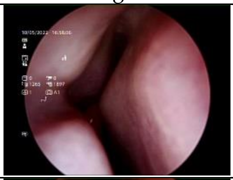 | 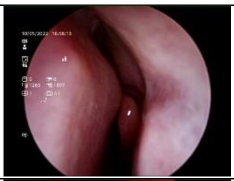 | Day 0 | 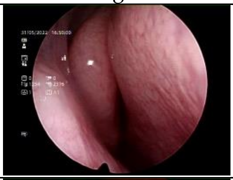 | 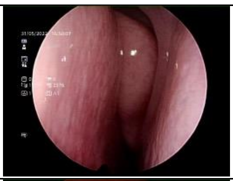 |
| Day 7 | 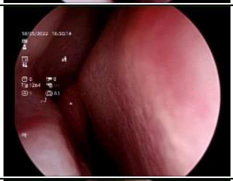 | 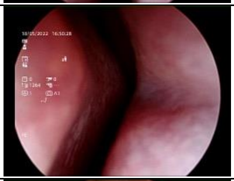 | Day 7 | 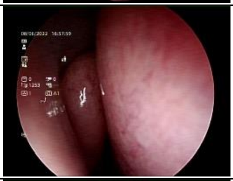 | 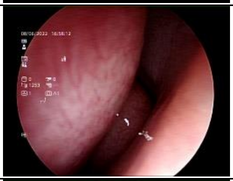 |
| Day 14 | 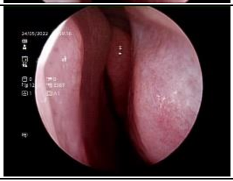 | 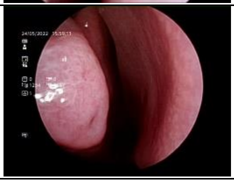 | Day 14 | 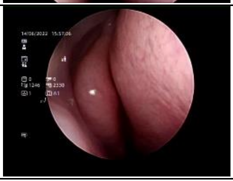 | 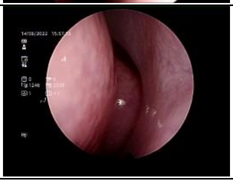 |

| | Nostril | | | Nostril | |
|---|---|---|---|---|---|
| C19 | Right | Left | C20 | Right | Left |
| Day 0 | 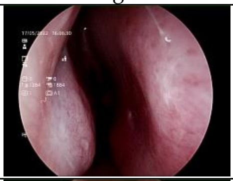 | 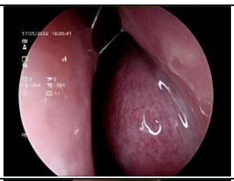 | Day 0 | 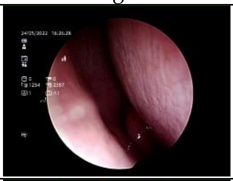 | 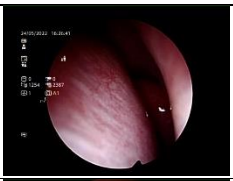 |
| Day 7 | 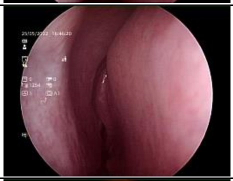 | 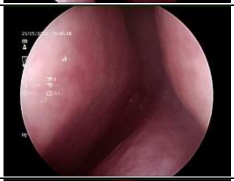 | Day 7 | 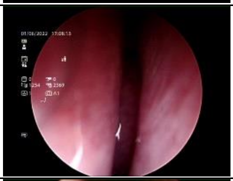 | 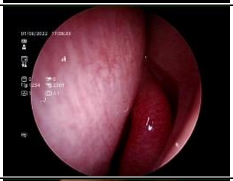 |
| Day 14 | 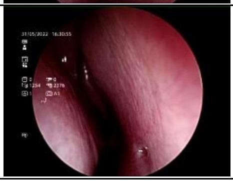 | 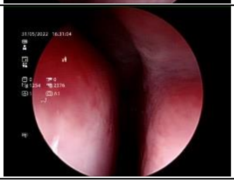 | Day 14 | 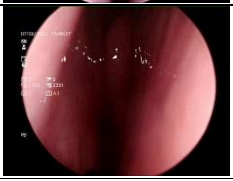 | 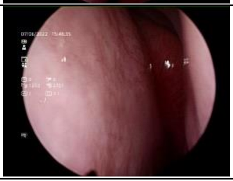 |
| C21 | Right | Left | C22 | Right | Left |
| Day 0 | 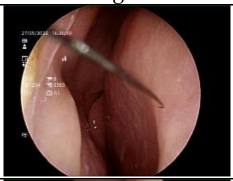 | 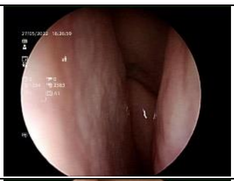 | Day 0 | 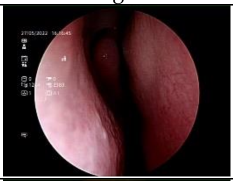 | 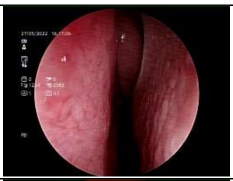 |
| Day 7 | 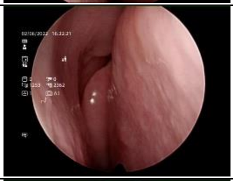 | 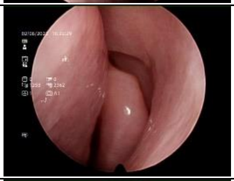 | Day 7 | 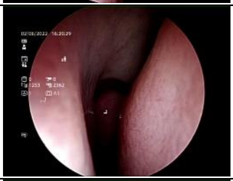 | 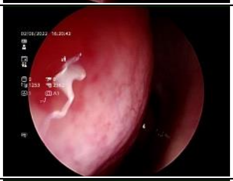 |
| Day 14 | 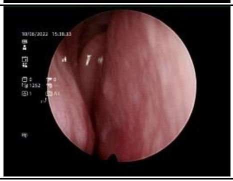 | 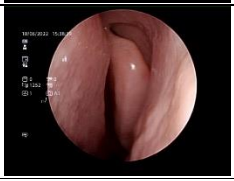 | Day 14 | 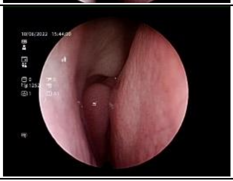 | 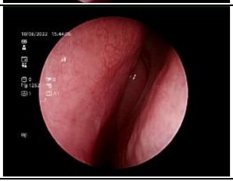 |
| C23 | Right | Left | C24 | Right | Left |
| Day 0 | 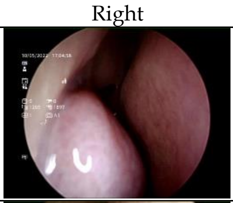 | 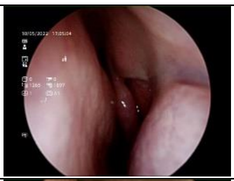 | Day 0 | 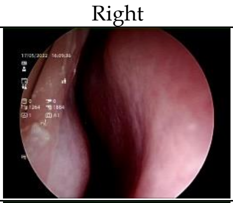 | 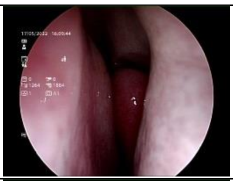 |
| Day 7 | 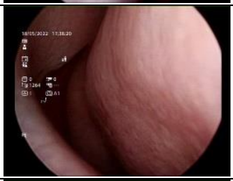 | 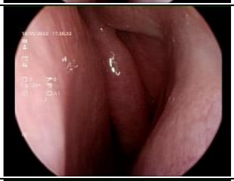 | Day 7 | 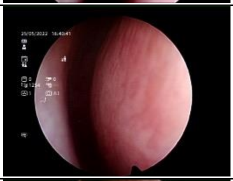 | 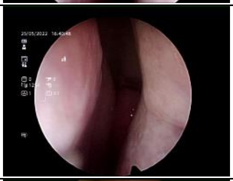 |
| Day 14 | 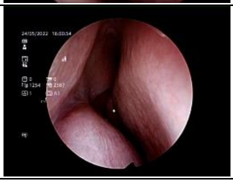 | 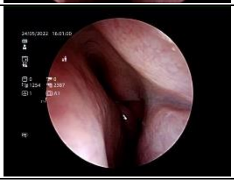 | Day 14 | 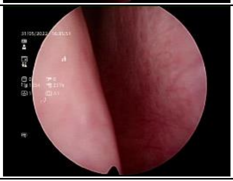 | 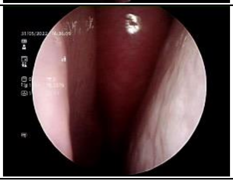 |

| | Nostril | | | Nostril | |
|---|---|---|---|---|---|
| C25 | Right | Left | C26 | Right | Left |
| Day 0 | 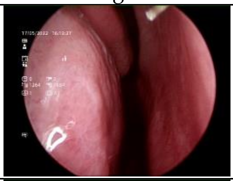 | 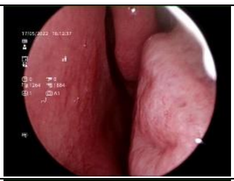 | Day 0 | 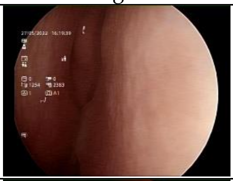 | 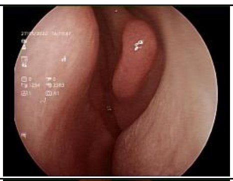 |
| Day 7 | 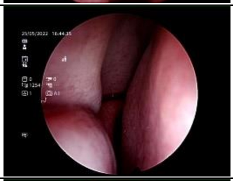 | 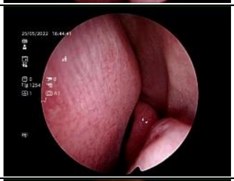 | Day 7 | 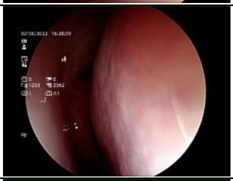 | 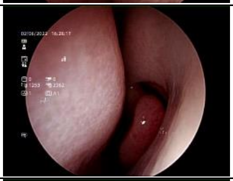 |
| Day 14 | 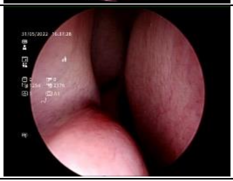 | 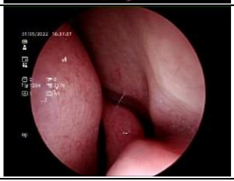 | Day 14 | 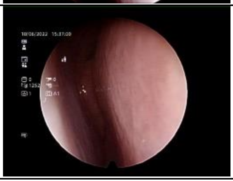 | 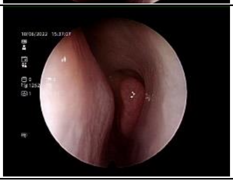 |
| C27 | Right | Left | | | |
| Day 0 | 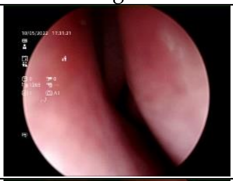 | 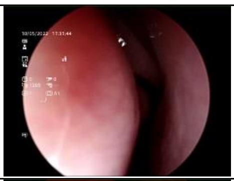 | | | |
| Day 7 | 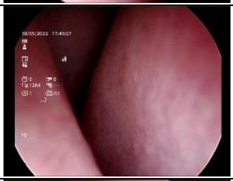 | 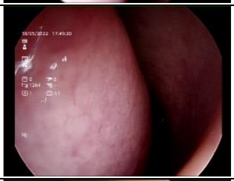 | | | |
| Day 14 | 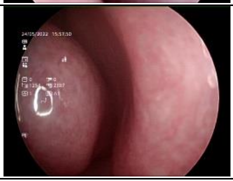 | 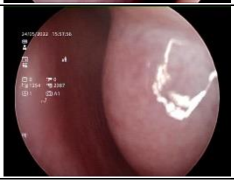 | | | |

#### Placebo group (n = 9)

| | Nostril | | | Nostril | |
|---|---|---|---|---|---|
| P1 | Right | Left | P2 | Right | Left |
| Day 0 | 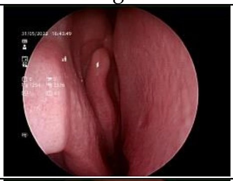 | 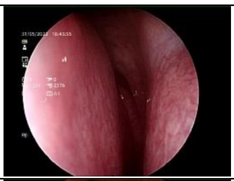 | Day 0 | 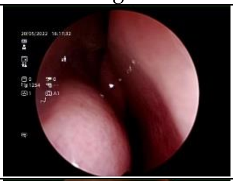 | 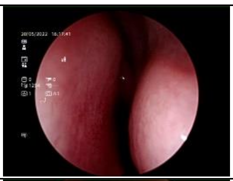 |
| Day 7 | 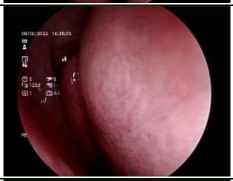 | 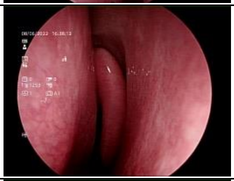 | Day 7 | 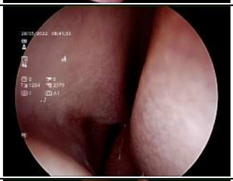 | 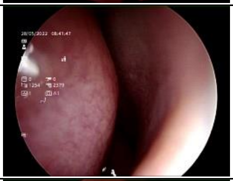 |
| Day 14 | 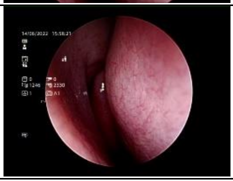 | 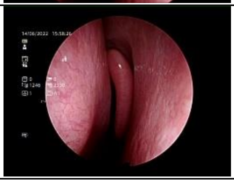 | Day 14 | 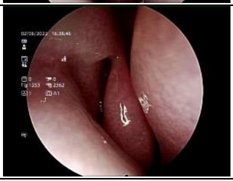 | 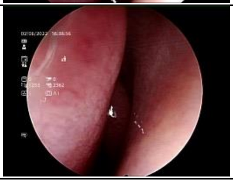 |
| P3 | Right | Left | P4 | Right | Left |
| Day 0 | 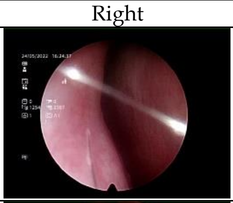 | 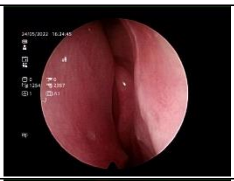 | Day 0 | 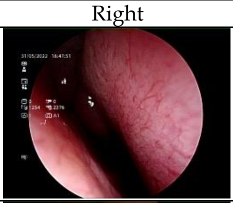 | 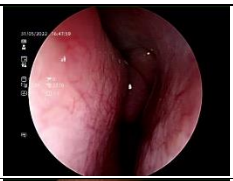 |
| Day 7 | 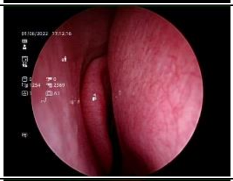 | 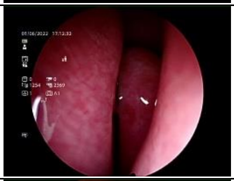 | Day 7 | 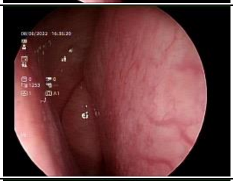 | 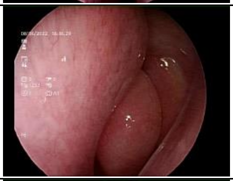 |
| Day 14 | 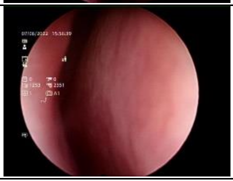 | 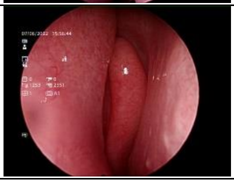 | Day 14 | 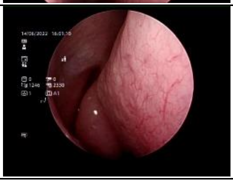 | 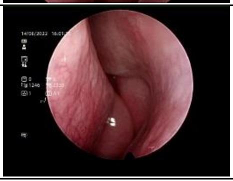 |
| P5 | Right | Left | P6 | Right | Left |
| Day 0 | 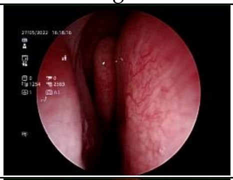 | 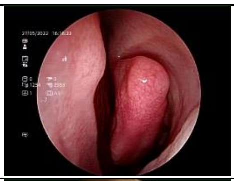 | Day 0 | 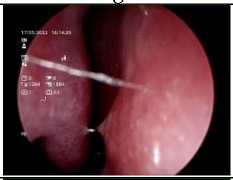 | 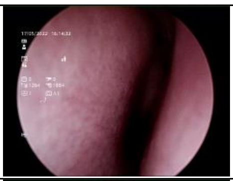 |
| Day 7 | 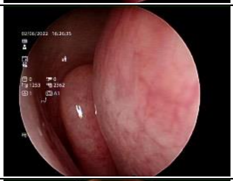 | 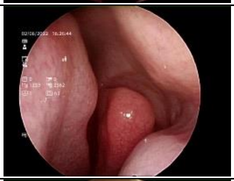 | Day 7 | 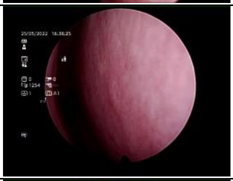 | 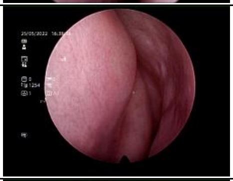 |
| Day 14 | 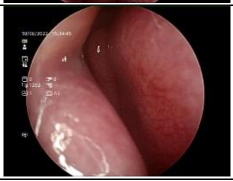 | 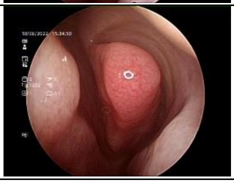 | Day 14 | 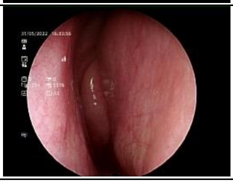 | 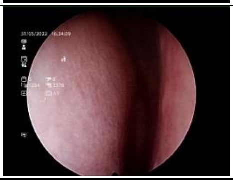 |

| | Nostril | | | Nostril | |
|---|---|---|---|---|---|
| P7 | Right | Left | P8 | Right | Left |
| Day 0 | 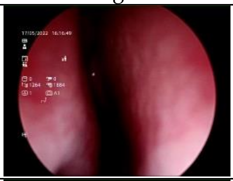 | 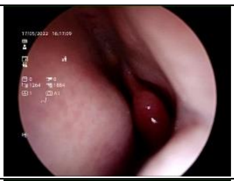 | Day 0 | 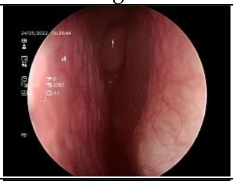 | 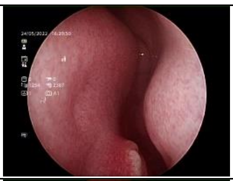 |
| Day 7 | 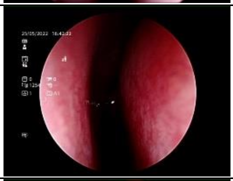 | 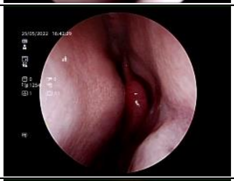 | Day 7 | 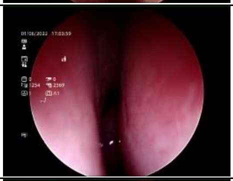 | 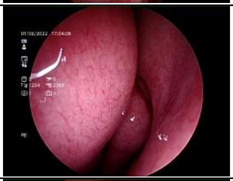 |
| Day 14 | 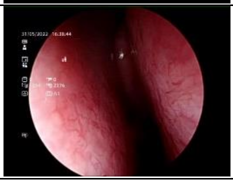 | 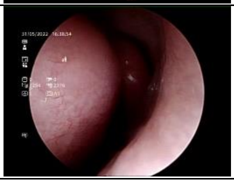 | Day 14 | 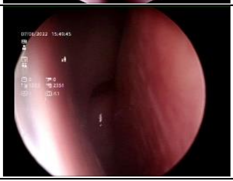 | 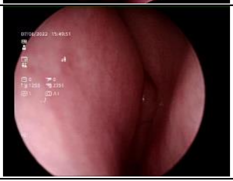 |

| P9 | Right | Left |
|---|---|---|
| Day 0 | 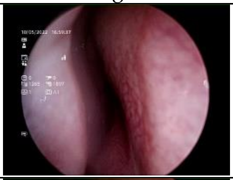 | 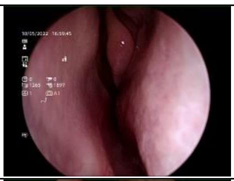 |
| Day 7 | 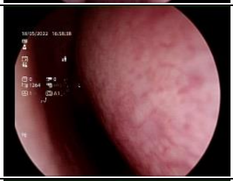 | 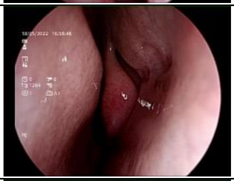 |
| Day 14 | 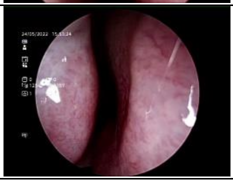 | 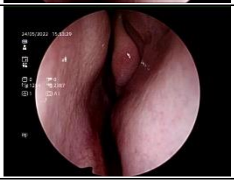 |
