## Supplemental Data S3 for "A randomized, placebo-controlled trial of a nasal spray solution containing broadly potent neutralizing antibodies against SARS-CoV-2 variants in healthy volunteers"

### Detailed statistical report of the intranasal SARS-CoV-2 inhibitory effects of COVITRAP™

| VOCs | Product | Timepoint of nasal fluid sample collection in relation to the study product application | *n* | Median of % inhibition (IQR25%-IQR75%) | 95% CI of median | Wilcoxon test (*P* value) |
|---|---|---|---|---|---|---|
| Ancestral | Placebo | Before | 4 | 7.69 (1.34 - 67.38) | 0 – 86.51 | 0.1875 |
| | | Immediately after | 4 | 14.35 (3.25 - 75.39) | 0 – 95.27 | |
| | COVITRAP™ | Before | 13 | 12.41 (3.56 - 27.39) | 0 – 33.63 | 1.2207E-4 |
| | | Immediately after | 13 | 97.58 (97.11 - 97.67) | 96.93 – 97.72 | |
| | Placebo | Before | 5 | 24.16 (6.48 - 94.45) | 0 – 97.73 | 0.1563 |
| | | 6 hours after | 5 | 46.68 (33.58 - 86.97) | 20.72 – 97.86 | |
| | COVITRAP™ | Before | 14 | 24.03 (12.2 - 34.9) | 4.06 – 38.36 | 6.1035E-05 |
| | | 6 hours after | 14 | 91.72 (76.2 - 97.04) | 66.43 – 97.29 | |
| Delta | Placebo | Before | 4 | 2.98 (2.25 - 49.94) | 2.13 – 65.46 | 0.3125 |
| | | Immediately after | 4 | 5.38 (0.53 - 65.7) | 0 – 84.73 | |
| | COVITRAP™ | Before | 13 | 8.02 (1.35 - 30.07) | 0.83 – 35.06 | 1.2207E-4 |
| | | Immediately after | 13 | 97.44 (96.88 - 97.57) | 96.61 – 97.62 | |
| | Placebo | Before | 5 | 35.01 (6.96 - 86.66) | 0 – 96.83 | 0.0938 |
| | | 6 hours after | 5 | 48.4 (23.64 - 86.17) | 23.08 – 97.63 | |
| | COVITRAP™ | Before | 14 | 21.53 (3.979 - 23.23) | 0 – 24.04 | 6.1035E-05 |
| | | 6 hours after | 14 | 88.67 (73.01 - 96.31) | 70.83 – 96.92 | |
| Omicron BA.2 | Placebo | Before | 4 | 2.16 (0 - 33.58) | 0 – 43.33 | 0.1250 |
| | | Immediately after | 4 | 12.5 (2.39 - 50.35) | 0 – 61.98 | |
| | COVITRAP™ | Before | 13 | 3.64 (0 - 10.99) | 0 – 11.58 | 1.2207E-4 |
| | | Immediately after | 13 | 94.65 (92.43 - 95.54) | 91.69 – 95.74 | |
| | Placebo | Before | 5 | 13.63 (3.14 - 50.38) | 0 – 70.69 | 0.0625 |
| | | 6 hours after | 5 | 17.77 (13.32 - 59.87) | 10.35 – 81.63 | |
| | COVITRAP™ | Before | 14 | 8.59 (0.09 - 15.79) | 0.04 – 16.78 | 6.1035E-05 |
| | | 6 hours after | 14 | 70.6 (54.83 - 85.09) | 48.33 – 88.95 | |
